# Accurate diagnosis of atopic dermatitis by applying random forest and neural networks with transcriptomic data

**DOI:** 10.1101/2022.04.04.22273382

**Authors:** Weixin Zhou, Aimin Li, Caiyun Zhang, Yongtao Chen, Zifan Li, Ying Lin

## Abstract

Atopic dermatitis (AD) is one of the most common inflammatory skin diseases. But the great heterogeneity of AD makes it difficult to design an accurate diagnostic pipeline based on traditional diagnostic methods. In other words, the AD diagnosis has suffered from an inaccurate bottleneck. Thus, it is necessary to develop a novel and accurate diagnostic model to supplement existing methods. The recent development of advanced gene sequencing technologies enables potential in accurate AD diagnosis. Inspired by this, we developed an accurate AD diagnosis based on transcriptomic data in skin tissue. Using these data of 149 subjects, including AD patients and healthy controls, from Gene Expression Omnibus (GEO) database, we screened differentially expressed genes (DEGs) of AD and identified six critical genes (PPP4R1, SERPINB4, S100A7, S100A9, BTC, and GALNT6) by random forest classifier. In a follow-up study of these genes, we constructed a neural network model (average AUC=0.943) to automatically distinguish subjects with AD from healthy controls. Among these critical genes, we found that PPP4R1 and GALNT6 had never been reported to be associated with AD. Although further replications in other cohorts are needed, our findings suggest that these genes may be developed into useful biomarkers of AD diagnosis and may provide invaluable clues or perspectives for further researches on the pathogenesis of AD.

## Introduction

Atopic dermatitis (AD) is one of the most prevalent inflammatory skin diseases, affecting about 15% of children and 7% of adults in the United States^1,2^. As reported by the WHO Burden of Disease, AD ranks the first with respect to the Global Burden of Skin Diseases (GBSD)^3^. Specifically, in addition to suffering AD-associated discomfort in the skin, when compared with healthy individuals, patients with AD have shown to have a higher risk of psychosocial comorbidities, such as depression, anxiety, sleep disorders and suicidal ideation, which brings about colossal economics cost and health threats to human society^4^. Formally, AD is a type of highly heterogeneous disease with different clinical phenotypes in terms of clinical features, severity, and course^5^. Elevation in total serum IgE level, eosinophilia, and allergen-specific IgE are the primary laboratory abnormalities of AD^6^. The pathophysiology is complex and involves a strong genetic predisposition, skin barrier disruption, T-cell driven inflammation, and microbial dysbiosis^7^. Currently, the therapeutic approaches remain limited, leaving difficulties in completely controlling AD in most patients^8^.

Due to the great heterogeneity mentioned above, it is difficult to define AD without its determinant laboratory biomarkers. The current AD clinical diagnostic rate is only 34.5%^9^. This phenomenon is due to the fact that there are no uniform standard diagnostic criteria that cover the entire spectrum of AD patients, and an experienced dermatologist’s assessment is considered the gold standard^10^. Traditional diagnostic criteria have been proposed to aid diagnosis mainly based on clinical manifestations and medical history. Among such criteria, the sensitivities of Hanifin and Rajka criteria^11^, Williams criteria^12–14^ and JDA criteria^15^ are 48.2%, 32.7%, 79.4% respectively in a hospital-based study^9^. The absence of uniform and accurate diagnostic criteria for AD limits the interpretability and comparability of clinical trials^16^. Moreover, it is often challenging to diagnose AD in adults who did not develop this disease in their childhood^17^.

Recently, the rapid development of second-generation sequencing technologies has facilitated the identification of many disease-related marker genes, including AD-related genes; for instance, Filaggrin (FLG) shows potential for screening and prognosis in AD^18^; CCL17 shows the highest correlation with AD severity^19^; GZMB, CXCL1, and CD274 show diagnostic value in skin tissue^20^. Furthermore, the gene expression data are increasingly used to establish diagnostic models of diseases with machine learning, especially random forest and artificial neural networks; for instance, heart failure, ulcerative colitis and polycystic ovary syndrome diagnostic models show excellent performance in accurate diagnosis^21–23^. However, there are currently few studies on prediction analysis using random forest and artificial neural networks based on the skin transcriptome in AD.

In conclusion, it is a challenge to diagnose AD with certainty because subjective clinical observations may not fully appreciate the variable clinical features. Therefore, it will have a potentially high impact on improving the accuracy of AD diagnosis to develop an objective and reliable biomarker for accurate AD diagnosis, especially in adults. In this study, based on the transcriptome of skin tissue, we build a classifier for accurate diagnosis of AD by integrating random forest and artificial neural networks. Firstly, we screen the Gene Expression Omnibus(GEO) database for differentially expressed genes (DEGs) between AD skin lesion samples and normal skin samples from healthy individuals. Then, based on these DEGs, we employ random forest and artificial neural network to select critical genes and calculate the weights of these genes, thus construct a genetic diagnostic model of AD. Finally, we apply this diagnostic model in other test sets to verify the efficiency of the designed neural-networks-based diagnostic pipeline. The flow chart is shown in Fig. 1.

**Figure 1.**
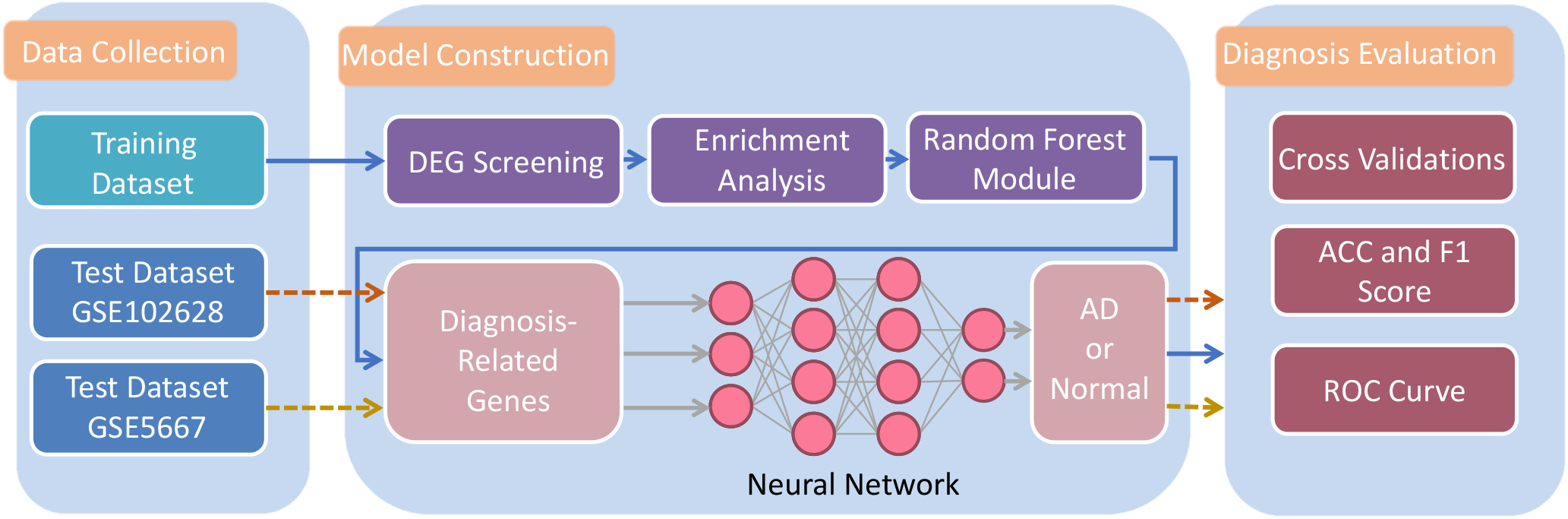
The flow chart in the study.

**Figure 2.**
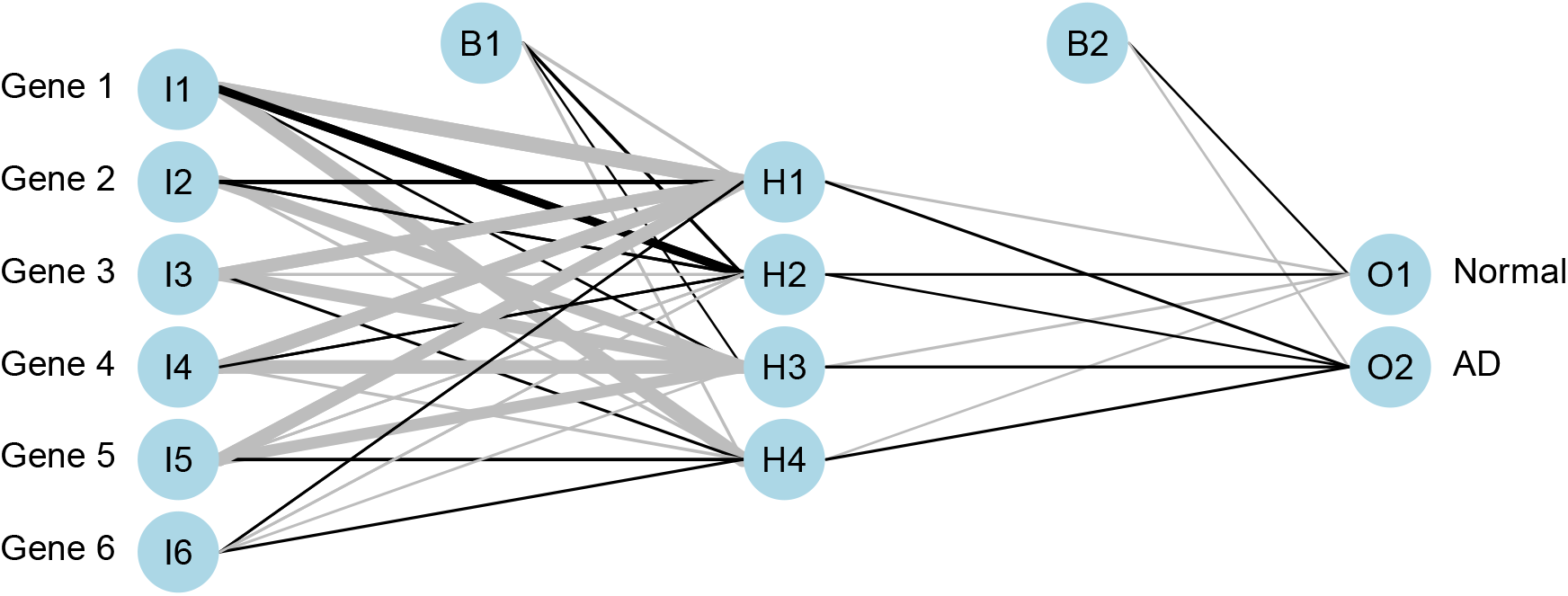
The construction of the neural network.

## Materials and Methods

### Public data collection

Five microarray datasets and two RNA-seq datasets were collected from the public Gene Expression Omnibus (GEO) database (https://www.ncbi.nlm.nih.gov/geo/). The detailed information of the collected datasets has been shown in Table 1. In order to obtain one training dataset with a relatively large sample size, five datasets (GSE6012, GSE32924, GSE60709, GSE121212, GSE153007) were combined and the batch effects were removed with R package “ *sva*” ^24^. Meanwhile, GSE102628 and GSE5667 were used as test datasets respectively. Only AD lesional tissue and normal tissue samples were chosen from the training dataset for further analysis. One test dataset (GSE5667) includes AD lesional tissue and normal tissue samples, while the other test dataset (GSE102628) includes AD non-lesional tissue and normal tissue samples. The patient information in each dataset has been shown in Supplementary Table S1.

**Table 1.** Detailed information of the collected GEO datasets.

| Dataset | Total samples | Normal Samples | AD Samples | Data Type | Country | Platform |
| --- | --- | --- | --- | --- | --- | --- |
| GSE6012 | 20 | 10 | 10 | Microarray | Sweden | GPL96 |
| GSE32924 | 21 | 8 | 13 | Microarray | USA | GPL570 |
| GSE60709 | 26 | 14 | 12 | Microarray | Germany | GPL6974 |
| GSE121212 | 65 | 38 | 27 | RNA-seq | USA | GPL16791 |
| GSE153007 | 17 | 5 | 12 | Microarray | USA | GPL6480 |
| GSE102628 | 19 | 9 | 10 | RNA-seq | Estonia | GPL20148 |
| GSE5667 | 11 | 5 | 6 | Microarray | USA | GPL96 |

### DEGs Screening

The training dataset containing 75 cases of AD and 74 cases of healthy control was used for differentially expressed genes (DEGs) analysis. The R package “ *limma*” was used to screen DEGs between the AD and control samples by the classical Bayesian method with adjusted *P <* 0.05 and |*log*2*FoldChange*| *>* 1^25^. DEGs screening was visualized by volcano plot and heat map^26,27^.

### Enrichment Analyses of DEGs

To determine functions of the identified AD-related DEGs, we performed Disease Ontology (DO), Gene Ontology (GO) and Kyoto Encyclopedia of Genes and Genomes (KEGG) pathway enrichment analyses, using R packages “ *clusterProfiler*” and “ *DOSE*” with *Q <* 0.05^28,29^. The GO analysis included biological process (BP), cellular component (CC) and molecular function (MF) categories. GObubble plots and GOChord plots were conducted with R package “ *GOplot*” to illustrate the functional analysis data^30^.

### Diagnosis-Related Gene Identification with Random Forest

As it is unlikely that all the identified DEGs are strongly associated with the occurrence of AD, it is necessary to further extract critical genes from these DEGs, with benefits from both accurate diagnosis and lower classification calculation complexity. Since the Mean Decrease Gini (MDG) method^31^ has shown superiority in screening important genes^21–23,32^, we leveraged it in the random forest classifier (RFC) to identify critical genes from DEGs.

### Parameters Selection

There are two important parameters that will significantly affect the performance of RFC implementation; one of them is called *mytry*, which represents the number of variables for the decision trees in the nodes; the other is denoted as *ntree*, which is the number of decision trees constructing the random forest model. For the optimal parameters setup, we first implemented a recurrent random forest classification for each possible value of *mytry* and calculated the average error rate. The value that enabled the minimal average error rate was selected as the optimal *mytry*. Then, we fixed *mytry* to this optimal value and plotted a figure regarding the relationship between *ntree* and the model error rate. The best *ntree* was finally selected based on a comprehensive consideration of model proficiency, model complexity, and model robustness.

### MDG Score Ranking

Based on the above-chosen parameters, we formulated the RFC model. During this process, the MDG score was also output as a default option. The MDG score is a metric that measures how much each input variable contributes to the homogeneity of the nodes and leaves in the generated RFC. Specifically, the higher its MDG score, the more influential the variable is in the model. In our study, we obtained the MDG score of each DEG by adopting R package “ *random forest*” ^33^. We accomplished feature extraction by ranking the DEGs in decreasing order of MDG scores and selecting the top *n*_*gene*_ genes as our candidate diagnosis-related genes.

### neural-networks-based AD Classifier

Once we screened out the most important genes, we next sought to construct a neural network model to automatically distinguish AD patients from normal people. Particularly, the construction of our designed neural-networks-based AD diagnostic model consists of several critical steps, which will be elaborated in the following.

### Normalizing

The inputs of the designed pipeline are the expressions with regard to the selected critical genes of each subject in the collected datasets. Denote the *j*^th^ critical gene expressions of the *i*^th^ sample in the collected dataset as *x*_*i, j*_ and define the average expression of the *j*^the^ diagnosis-related gene as 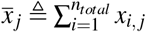, the normalization process is given as

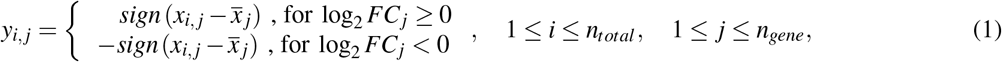

where *y*_*i, j*_ is the normalized data, *n*_*total*_ represents the number of samples, and log2 *FC*_*j*_ denotes the *log*2*FoldChange* value of the *j*^th^ critical gene that has been calculated in the previous subsection **DEGs Screening**, and the function *sign* (·) is defined as

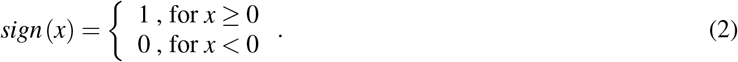

The above normalization process forms a threshold-segmentation structure. In particular, if the *log*2*FoldChange* value of a gene is greater than 0, the gene expression will exert a positive impact on the occurrence of AD, a value greater than the average gene expression is normalized to 1, and the one lower than the average gene expression is normalized to 0; otherwise, the gene expression will exert a negative impact on the occurrence of AD, and the value is normalized in an opposite way. The implementation of thresholding and normalizing mainly brings about two advantages: *i*) the scaled gene expressions are transformed to positive numbers; *ii*) the threshold-segmentation amplifies the effect of up-regulated genes on the pathogenesis, and thus enables improvement in the diagnosis performance of the classifier. Here both the training and test datasets are normalized together.

### Neural Networks Constructing

We constructed the neural network by utilizing R packages “ *neuralnet*” and “ *Neu-ralNetTools*” ^34^. The constructed neural network consists of an input layer with *n*_*gene*_ = 6 neurons, a hidden layer with four neurons, and an output layer with two neurons. The details of the neural network structure were demonstrated in Figure 6 (**a**). The training process was conducted according to the normalized training data matrix denoted by 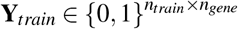, where *n*_*train*_ is the size of the training dataset, *n*_*gene*_ is the number of selected diagnosis-related genes. The training dataset is marked with a label vector 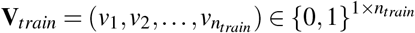 with *v*_*i*_ = 1 if the *i*^th^ sample is from AD lesional tissue and *v*_*i*_ = 0 if the *i*^th^ sample is from normal tissue. The goal of the training process is to obtain the weighted matrix **W** and the bias vector **B** at each layer, so that the output of the neural network turns closer to the label vector **V**_*train*_. Details of weighted parameters are shown in Supplementary Table S6.

### Performance Metric

The results of the binary classification can be directly visualized by a 2 × 2 matrix, which is known as the confusion matrix (Table 2).

**Table 2.** An example of the confusion matrix.

| Diagnosis Experiments |  | Predicted Class |  |
| --- | --- | --- | --- |
|  |  | AD | Normal |
| Actual Class | AD | TP<br>(True Positive) | FN<br>(False Negative) |
|  | Normal | FP<br>(False Positive) | TN<br>(True Negative) |

From a confusion matrix, several critical performance metrics can be obtained straightforwardly, which are True Positive Percentage (TPP), False Positive percentage (FPP), Accuracy (ACC), and F1 Score. We elaborate on these performance metrics as follows.

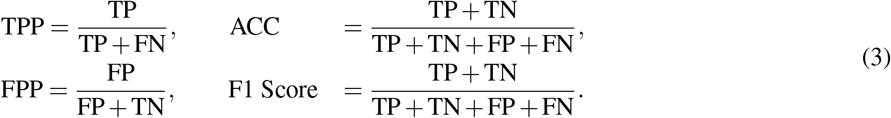

We mainly focused on ACC and F1 score in this paper to evaluate the performance of the neural-networks-based AD classifier. The ACC is defined as the proportion of correctly classified samples to total samples. In contrast, the F1 score can provide a more integrated classifier performance evaluation, especially when the dataset is imbalanced. Both ACC and F1 score are real numbers ranging from 0 to 1. The closer the ACC (or F1 score) is to 1, the better the diagnostic model performs.

In addition to the aforementioned metrics, another integrated evaluation method is the Receiver Operating Characteristic (ROC) curve. The ROC curve sketches the relationship between TPP and FPP at various threshold settings. In such cases, the ROC curve not only focuses on a single classifier, but also broadens its evaluations to a series of classifiers that can be generated by different classifying threshold setups. Once we obtain the ROC curve, we can then calculate the area under the ROC curve (AUC). The AUC value also ranges from 0.5 to 1, where the value 1 represents the perfect and efficient diagnosis while 0.5 represents the inapplicable random diagnosis.

### Cross-Validations on the Training Dataset

To verify the robustness of our proposed model, we also conducted 6-fold cross-validations (CV) in this work. Specifically, the normalized training dataset **Y**_*train*_ is randomly divided into six uniform sub-matrices, one of which is regarded as a test set while the others are input to the neural network for the training purpose. This 6-fold CV process will be conducted six times, so that each of the six sub-matrices will serve as a test dataset once, and evaluation metrics such as ACC, F1 Score, ROC curve, and AUC will be obtained correspondingly.

### Validations on Test Dataset

Up to this point, we have acquired the weighted matrix and the bias vector in the neural network through the training process. The next step is to apply this diagnostic model in other test datasets, so as to verify the efficiency of the designed neural-networks-based diagnostic pipeline. During the validation process, the test matrix **Y**_*test*_ is input to the neural network to generate the predicted results. According to the predicted results, the ACC, F1 Score, ROC curve, and AUC can also be obtained.

## Results

### Identification of DEGs

A total of 288 DEGs were identified in the training dataset, consisting of 188 up-regulated and 100 down-regulated genes (Fig.3(**a**) and Supplementary Table S2). The heat map (Fig.3(**b**)) displays the expression level of each gene in different samples, showing that the overall expression level of DEGs differed greatly between AD lesional skin tissues and normal skin tissues.

**Figure 3.**
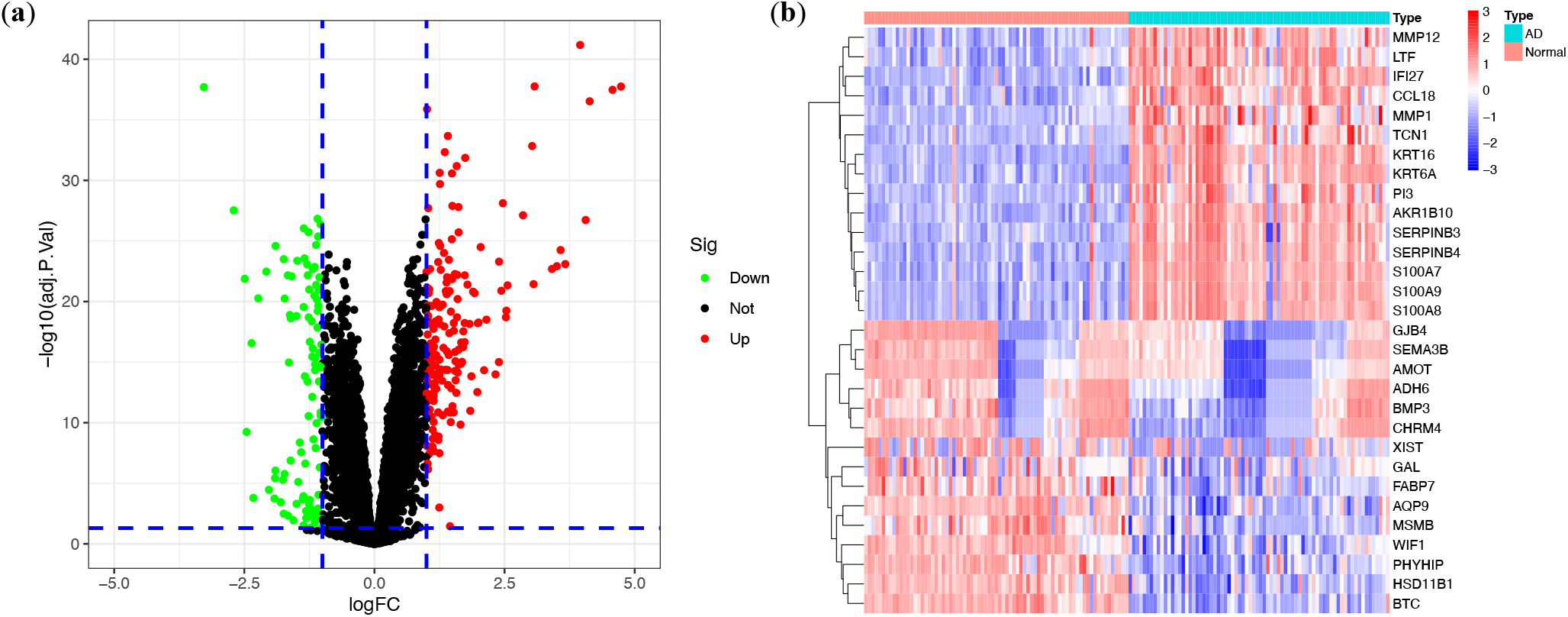
Analyses of DEGs in the training dataset.(**a**)Volcano plot of DEGs in lesional vs.normal tissues. Red dots represent up-regulated genes and green dots represent down-regulated genes. The black dots indicate the remaining stable genes. DEGs are defined as the cut-off criteria of |*log*2*FoldChange*| *>* 1 and adjusted *P <* 0.05. (**b**)Heat map of the top 15 up-regulated and down-regulated DEGs in lesional vs. normal tissues. Red represents DEGs with high expression, while blue represents DEGs with low expression.

### Functional Enrichment Analysis

To explore the potential related diseases and underlying mechanisms of the 288 DEGs, DO, GO, and KEGG enrichment analyses were conducted. GO enrichment analysis for BP category indicated that the DEGs were enriched in cytokine-mediated signaling pathway, response to virus and granulocyte migration; for CC category, tertiary granule lumen, cornified envelope and external side of plasma membrane were significantly enriched; for MF category, chemokine activity, serine-type endopeptidase activity and serine-type peptidase activity were significantly enriched (Fig.4 (**a**) and Supplementary Table S3). The KEGG analysis suggested that the DEGs were significantly enriched in Viral protein interaction with cytokine and cytokine receptor, cytokine-cytokine receptor interaction and IL-17 signaling pathway (Fig.4 (**c**) and Supplementary Table S4). The DO enrichment analysis showed that the DEGs were enriched in integumentary system disease, bacterial infectious disease, atopic dermatitis, etc. (Fig.4 (**e**) and Supplementary Table S5) Part of GO, KEGG and DO enriched terms and the significant DEGs involved were showed respectively in Fig.4 (**b**), Fig.4 (**d**) and Fig.4 (**f**).

**Figure 4.**
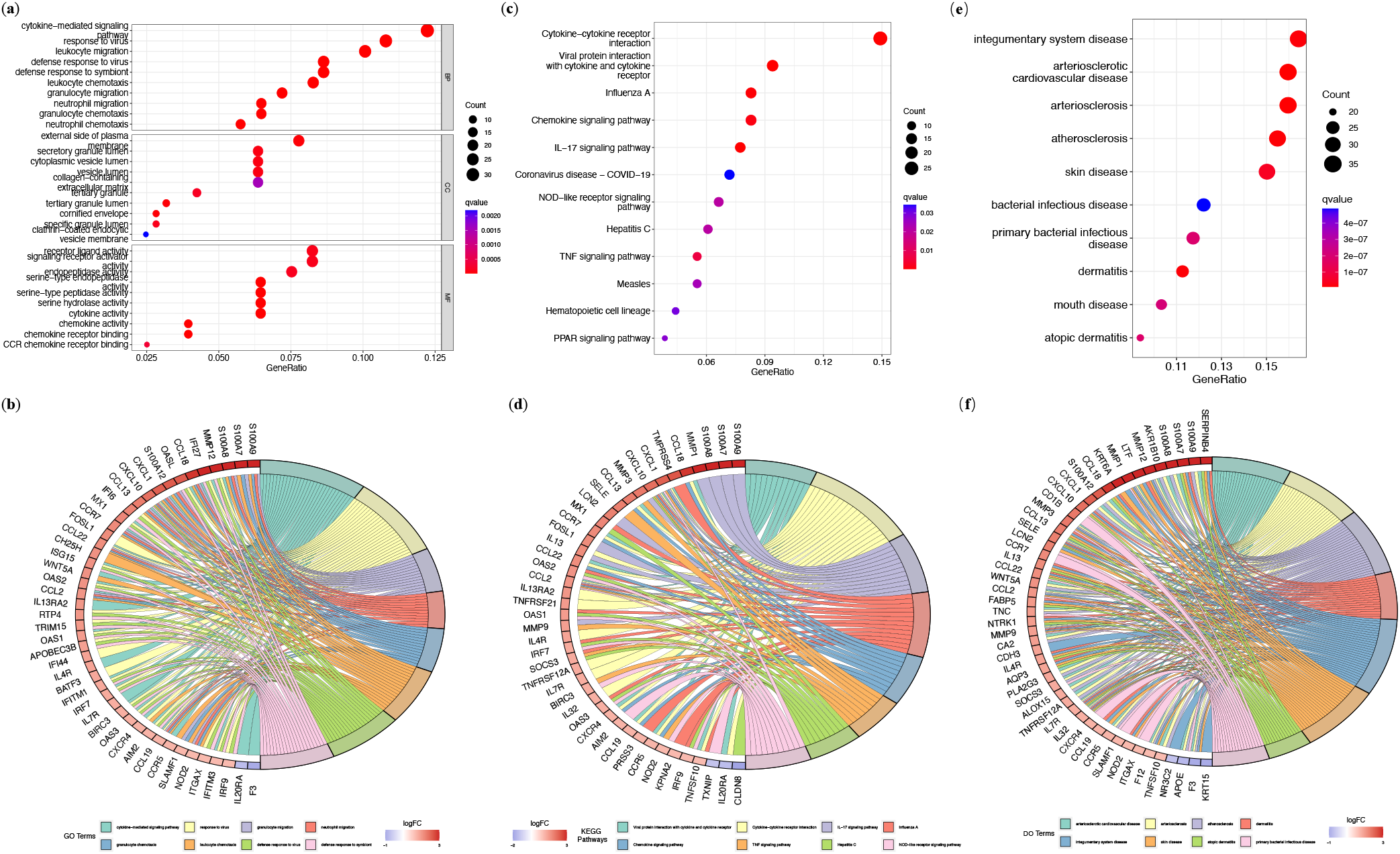
Enrichment Analyses of DEGs in AD. (**a**) Bubble plot showing significantly enriched GO terms included biological process (BP), cellular component (CC) and molecular function (MF) categories. (**b**) Chord plot of the top 8 enriched GO terms with minimum *Q* values. (**c**) Bubble plot of 12 enriched KEGG pathways. (**d**) Chord plot of the top 8 enriched KEGG pathways with minimum *Q* value. (**e**) Bubble plot of the top 10 enriched DO terms with minimum *Q* value. (**f**) Chord plot of the top 8 enriched DO terms with minimum *Q* value.

### The Diagnosis-Related Genes

Fig. 5 (**a**) and Fig. 5 (**b**) demonstrate the detailed variables selection results while constructing the random forest. In Fig. 5 (**a**), the scatter plot is carried out to show the relationship between the out-of-band error rate and *mytry*. As the red point in the figure exhibits the lowest error rate, we finally choose *mytry* = 17 for the best *mytry* setup. Fig. 5 (**b**) shows the relationship between *ntree* and the error rate. We set *n*_*tree*_ = 400 as the parameter of the RFC model because such a setting demonstrates superiority in both low error rate and stable system performance.

**Figure 5.**
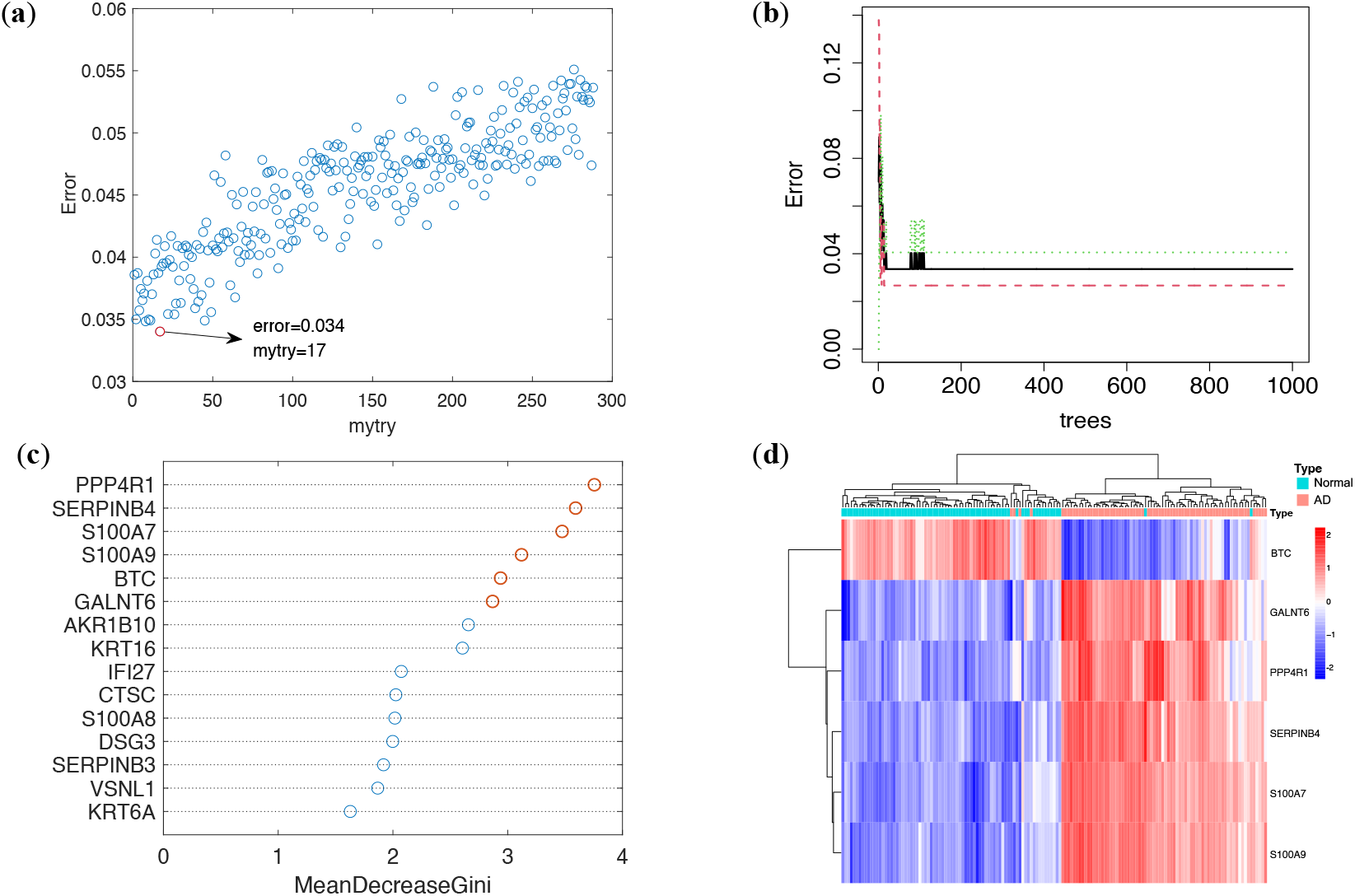
Diagnosis-related genes selection process. (**a**) Scatter plot regarding the relationship between *mytry* and the out-of-band error rate. (**b**) Error rate vs. *ntree*. (**c**) Results of the top 15 MDG scores. The red points indicate MDG scores of the selected diagnosis-related genes. (**d**) Heatmap of the selected diagnosis-related genes.

Fig. 5 (**c**) shows the top 15 genes in MDG scores, wherein the top 6 genes (i.e., PPP4R1, SERPINB4, S100A7, S100A9, BTC, and GALNT6) are selected as the diagnosis-related genes. In Figure. 5 (**d**), the heatmap is drawn to visually show the expression levels of each selected genes in different samples. From Fig. 5 (**d**), we can observe that the overall expression levels of the selected genes differed greatly between AD samples and normal samples, which demonstrates that the selected genes enable potential to be new biomarkers of AD, thus bringing about new insights in the diagnosis of AD.

### Six-Fold Cross-Validations

Cross-validation (CV) is a general-purpose method to evaluate whether a neural network is overfitted^35^. Therefore, we performed the six-fold CV to evaluate this particular characteristic of our AD-diagnosis-oriented model. To this end, the ROC curves together with the AUC, ACC and F1 Score are shown in this subsection.

Figure. 6 aims to display the robustness of our model by giving the ROC charts of each cross-validation. In addition, Table 3 demonstrates the specific ACC, F1 Score and AUC of each cross-validation. All these performance metrics are close to 1, verifying that the constructed neural network is not overfitted, offering superior robustness and generalization.

**Figure 6.**
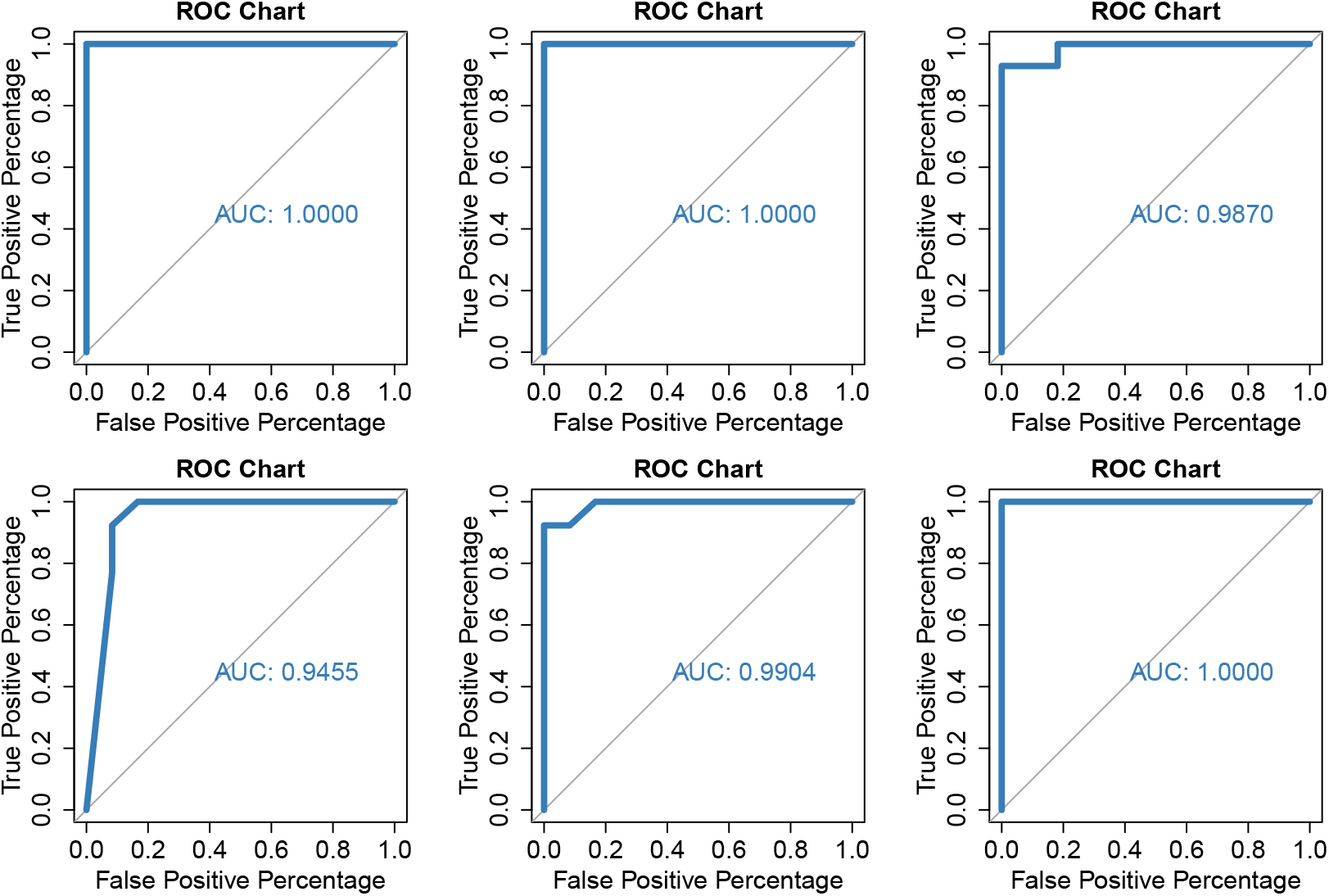
ROC curves of 6-fold cross-validations.

**Table 3.** ACC, F1 Score and AUC of the 6-fold cross-validations.

| Cross-Validations | ACC | F1 Score | AUC |
| --- | --- | --- | --- |
| CV1 | 1.000 | 1.000 | 1.000 |
| CV2 | 1.000 | 1.000 | 1.000 |
| CV3 | 0.960 | 0.962 | 0.987 |
| CV4 | 0.960 | 0.923 | 0.945 |
| CV5 | 0.958 | 0.960 | 0.990 |
| CV6 | 1.000 | 1.000 | 1.000 |

### Performance Evaluation

In an available research, CXCL1 and GZMB are reported as critical biomarkers in prediction of the incidence of AD^36^. As such, we conducted performance comparisons among the neural-networks-based diagnostic model, the CXCL1-based diagnostic model, and the GZMB-based diagnostic model. Diagnosis experiments were carried out on the training dataset and the independent test datasets (GSE5667 and GSE102628) respectively.

Figure. 7 shows the comparison of the ROC charts of the three diagnosis baselines. Evaluations were conducted on the training dataset, the GSE5667, and the GSE102628, respectively. In addition, Table 4 demonstrates the specific ACC, F1 Score and AUC to verify the strength of the neural-networks-based model. It shows that the proposed neural-networks-based diagnosis draws its strength from excellent accuracy and higher robustness, compared with the single-biomarker-based diagnosis.

**Figure 7.**
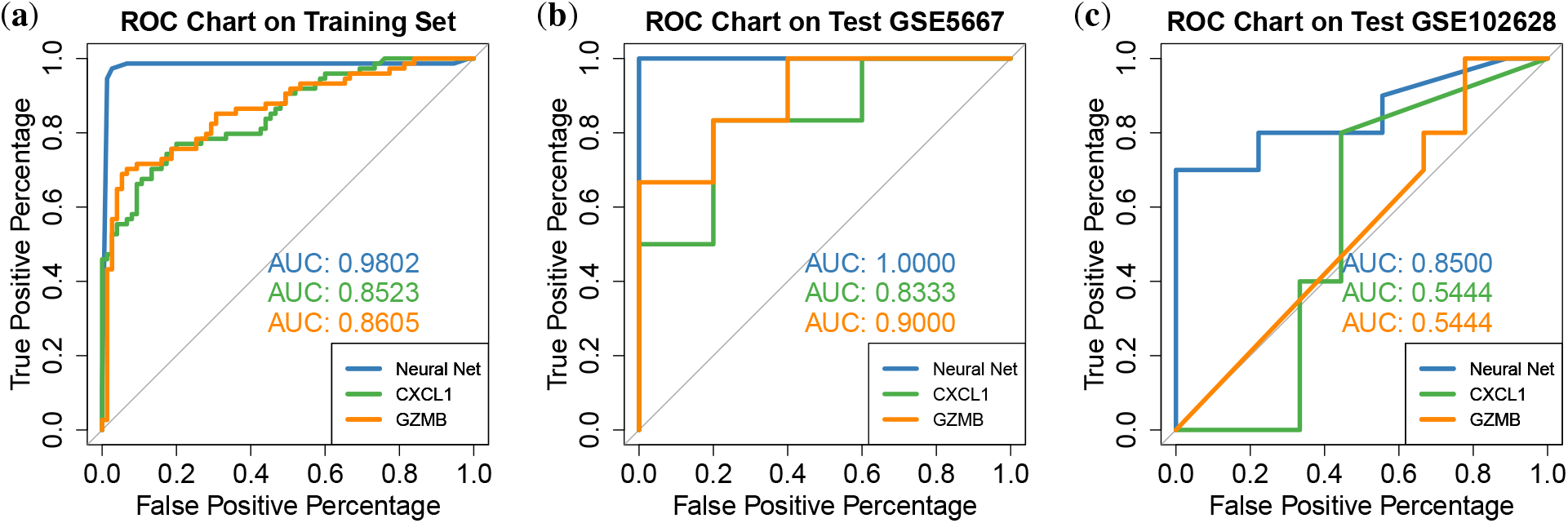
ROC curves on training dataset, test dataset GSE5667 and test dataset GSE102628.

**Table 4.** Evaluation results with regard to the ACC, F1 Score and AUC.

| Diagnosis Experiments | ACC | F1 Score | AUC |
| --- | --- | --- | --- |
| neural-networks-based Diagnosis on the Training Dataset | 0.973 | 0.972 | 0.980 |
| CXCL1-Based Diagnosis on the Training Dataset | 0.785 | 0.780 | 0.852 |
| GZMB-Based Diagnosis on the Training Dataset | 0.819 | 0.794 | 0.850 |
| neural-networks-based Diagnosis on Test Dataset GSE5667 | 1.000 | 1.000 | 1.000 |
| CXCL1-Based Diagnosis on Test Dataset GSE5667 | 0.878 | 0.833 | 0.833 |
| GZMB-Based Diagnosis on Test Dataset GSE5667 | 0.878 | 0.833 | 0.900 |
| neural-networks-based Diagnosis on Test Dataset GSE102628 | 0.842 | 0.823 | 0.85 |
| CXCL1-Based Diagnosis on Test Dataset GSE102628 | 0.684 | 0.727 | 0.544 |
| GZMB-Based Diagnosis on Test Dataset GSE102628 | 0.631 | 0.744 | 0.544 |

## Discussion

AD is a common but complex disorder characterized by increasingly recognized diverse clinical and molecular phenotypes which may be generated by gene-gene and gene-environment interactions^37^. Due to this diversity, clinical observations may not entirely appreciate skin lesions, posing enormous diagnostic and therapeutic challenges. Thus, it is imperative to explore reliable and objective biomarkers for AD diagnosis. The purpose of this study was to predict phenotypes of AD with gene expression data. We constructed an AD diagnostic classifier solely based on gene expression data obtained from GEO database. We integrated and took advantage of random forest and artificial neural networks to infer important candidate classification genes and calculate the weights of these genes. Our diagnostic model with 6 selected critical genes automatically classified AD with excellent AUC efficiency (up to 1.00). It accurately distinguished subjects with AD from healthy controls. Among the critical genes, we found that PPP4R1 and GALNT6 had never been reported to be associated with AD. In summary, our classifer represents better prediction performance than utilizing previously reported marker genes and provides important biological insights into the development of the biomarkers of this disease.

In GO enrichment analysis based on the AD-related DEGs, the following aspects were significantly enriched: 1) inflammation-related processes: cytokine-mediated, granulocyte migration, neutrophil migrationl, etc.; 2) immune response processes: lymphocyte chemotaxis, regulation of innate immune response, T cell migration, etc.; 3) bacterial response processes: defense response to bacterium, antimicrobial humoral immune response mediated by antimicrobial peptide, response to molecule of bacterial origin, etc.; 4) skin-related processes: skin development, epidermal cell differentiation, epidermal cell differentiation, etc.; these results represented that the over activation of inflammatory and immune responses and the dysfunction of epidermis development processes occupied an essential position in the development of AD. In addition, several immune-related pathways were significantly enriched in the KEGG pathway, such as cytokine-cytokine receptor interaction, IL-17 signaling pathway, chemokine signaling pathway. As primary mediators of the adaptive immune system, cytokines including IL-4, IL-5, IL-13, IL-31and IL-33 are important players in the Th2 immune response of AD^38^. IL-17, a key cytokine for activating macrophages and fibroblasts, can induce antimicrobial peptide expression, which may contribute to bacterial infection frequently seen in AD^39^. Interestingly, by executing the DO analysis, we observed that the DO analysis significantly enriched in not only atopic dermatitis but also artery diseases. This observation indicates that it may have some potential pathogenesis relationships between these two diseases. In practice, vascular inflammation in AD is linked to Th2 response and clinical severity that has been suggested previously^40^. Among the six critical genes, three up-regulated genes (S100A7, S100A9, SERPINB4) are involved in multiple immune processes, while one down-regulated gene (BTC) plays a prominent role in positive regulation of epidermal growth. S100A7 and S100A9, members of the S100 family, can exhibit antimicrobial activities and induce immunomodulatory activities in hyperproliferative skin diseases^41^. SERPINB4 has crucial effects on initiating early inflammatory response and can lead to chronic skin inflammation like AD^42^. BTC is one of the epidermal growth factors, playing important roles in skin morphogenesis, homeostasis, and repair^43^. It is worth noting that the function of PPP4R1 and GALNT6 on AD remains unclear, but these two genes are among the top 15 DEGs with minimum adjusted *P* value in AD (Supplementary Table S2). Therefore, these six genes can be used as promising biomarkers to establish a diagnostic model of AD.

To the best of our knowledge, this is the first attempt to apply random forest and neural network algorithms with gene expression profiles in AD diagnosis. Compared with the other two potential gene biomarkers reported previously for AD diagnosis^36^, the proposed neural-networks-based diagnostic model in our study has drawn its strength from more excellent accuracy, as well as higher robustness in the training set (AUC=0.9802) and two validation sets (AUC=1.0000 in GSE5667 and AUC=0.8500 in GSE102628 respectively). Our diagnostic model covered patients with AD of different races, severities, and disease courses, greatly increasing the universality of the diagnosis. It should be noted that GSE102628 is a special dataset different from other collected datasets. Specifically, the transcriptomic data of the AD samples in GSE102628 were obtained from their non-lesional skin tissues. In such a case, we can witness an evident performance loss if CXCL1 or GZMB was adopted sorely for the diagnosis of AD, i.e., the AUC of such a diagnostic model is close to 0.5. Nevertheless, the proposed neural-networks-based diagnostic model remains efficient on GSE102628, validating the generalization of the neural-networks-based model. Previously, several sets of diagnostic models of AD using omics data were established to explore the potential diagnostic biomarkers. Based on the transcriptomic data for host gene expression, 16S rRNA microbiome and metagenome shotgun data of the intestinal microbiota, Park, J et al. employed machine learning to construct a diagnostic model, the AUC of which was 0.715^44^. In a similar manner, AUC of another machine learning diagnostic model based on the transcriptome of gut epithelial colonocytes and gut microbiota data was 0.75^45^. Besides, a diagnostic model developed based on microbiome in serum extracellular vesicle analysis showed high accuracy (AUC=1.00), but without external validation. As we know, the accessibility of skin makes it the perfect tissue for the investigation of skin disease. With the emergence of minimally invasive methods, such as tape strips, access to skin sampling from both adults and children may become more convenient and easier in the near future^46^. Meanwhile, it might be more cost-effective to obtain transcriptomic data with the rapid development of second-generation sequencing technologies. Thus, with a high degree of predictive accuracy, our classifier is promising to assist clinicians to perform accurate AD diagnosis and inform clinical decision-making in the future.

There are also limitations in our study. We elaborate some of them here, which may facilitate future works. First, the neural network training process in this paper was conducted on a dataset with a sample size of 149, which may not be sufficient for the data-driven learning method, thus might cause overfitting and biases when training the neural-networks-based model. To verify the robustness of our proposed model, we applied 6-fold cross-validations and two independent validation datasets, whose performance showed that we successfully controlled the biases and overfitting of our diagnostic model. Second, the number of the validation datasets in this paper was also limited. In this regard, it may be valuable to generalize our diagnostic model into more AD-diagnosis scenarios, so as to further test the effectiveness of the diagnostic model. Third, since the ages of our subjects range from 16 to 81, there is a possibility that the results may differ in children. Therefore, in order to validate and improve the neural-networks-based model and evaluate its performance more accurately, further studies in larger sample sizes and independent cohorts are needed. Although this is a compromising strategy in the situation of limited sample size, our model has shown an excellent classification performance. A diagnostic model for AD patients of all ages still needs to be constructed with more convincing datasets in the future.

In this study, we offered pieces of evidence that the transcriptome in skin tissue might be a critical factor in disease diagnosis. Through transcriptome utilization, objective and reliable diagnostic biomarkers could be attained. The 6 critical genes (PPP4R1, SERPINB4, S100A7, S100A9, BTC, and GALNT6) in our diagnostic model may be developed into valuable biomarkers of AD diagnosis and may provide invaluable clues or perspectives for further researches on the pathogenesis of AD.

## Data Availability

All data produced are available online at https://www.ncbi.nlm.nih.gov/geo/

## Author contributions statement

W.Z., Y.L. and Y.C. conceived the study and designed the experiments, W.Z. and A.L. conducted the experiments and analyzed the results, W.Z., A.L., C.Z. and Z.L. drafted the manuscript. All authors reviewed the manuscript.

## Data availability

The data supporting the findings of this study are included within the article.

