## Supplemental Table 1 for "Accurate diagnosis of atopic dermatitis by applying random forest and neural networks with transcriptomic data"

**Supplementary Tables**

**Table S1.** The patient information in each dataset

| Dataset | Patient Age(years) | Control Age | Patient Severity | Race |
| --- | --- | --- | --- | --- |
| GSE6012 | 40.3 (21–50) | 43(25-50) | Scorad 42.6 (20.6–60) | Caucasian |
| GSE32924 | 38 (16–81) | - | Scorad 60 (28 to 97.5) | - |
| GSE60709 | 38.50(20-62) | 27.00(22-69) | moderate to severe | - |
| GSE121212 | 34.07+/-10.96 | 32.63+/-11.64 | Scorad 31.11+/-10.96 | - |
| GSE153007 | 36.92(19-65) | 51.80(44-58) | Mild to severe | - |
| GSE102628 | 25.70(18-38) | 46(24-64) | EASI 11.90+/-3.50 | - |
| GSE5667 | 31.67（18-44） | 28.60（22-35） | mild to severe | White,Asian, Hispanic, Black |

**Table S2.** Differentially expressed genes in lesional skin tissues and non-lesional skin tissues

| id | logFC | adj.P.Val |
| --- | --- | --- |
| KRT16 | 3.95172756 | 6.66E-42 |
| IFI27 | 3.07663815 | 1.76E-38 |
| S100A7 | 4.73592367 | 1.80E-38 |
| S100A9 | 5.11992681 | 1.99E-38 |
| BTC | -3.276341 | 1.99E-38 |
| S100A8 | 4.57474462 | 3.34E-38 |
| AKR1B10 | 4.13510517 | 2.95E-37 |
| PPP4R1 | 1.01014026 | 1.33E-36 |
| SERPINB4 | 5.72484789 | 7.55E-35 |
| RAB31 | 1.41073316 | 2.15E-34 |
| KRT6A | 3.03006338 | 1.46E-33 |
| IL4R | 1.35181851 | 4.68E-33 |
| GALNT6 | 1.74346781 | 1.39E-32 |
| S100A2 | 1.5797753 | 6.58E-32 |
| SCO2 | 1.25354421 | 2.38E-31 |
| DSG3 | 1.48930422 | 2.68E-31 |
| VSNL1 | 1.25917042 | 2.00E-30 |
| TMPRSS4 | 2.46791909 | 7.59E-29 |
| CTSC | 1.49729142 | 1.27E-28 |
| WNT5A | 1.6145374 | 1.60E-28 |
| KPNA2 | 1.03326955 | 1.96E-28 |
| HSD11B1 | -2.7031166 | 2.97E-28 |
| CCL18 | 2.85431631 | 7.53E-28 |
| GPLD1 | -1.0942355 | 1.46E-27 |
| SERPINB3 | 4.05763803 | 1.85E-27 |
| SCCPDH | -1.0366754 | 4.16E-27 |
| ZSCAN18 | -1.3576179 | 9.10E-27 |
| BCAR3 | -1.2677358 | 1.83E-26 |
| DSC2 | 1.61729142 | 1.93E-26 |
| FAM134B | -1.0817638 | 4.33E-26 |
| RAI14 | -1.0713815 | 4.33E-26 |
| TNC | 1.48646931 | 7.21E-26 |
| MPZL2 | 1.23740046 | 1.47E-25 |
| RHOBTB3 | -1.1223192 | 2.15E-25 |
| PHYHIP | -1.8989637 | 2.54E-25 |
| IRF7 | 1.26443382 | 2.56E-25 |
| ABCG4 | 2.04024558 | 3.20E-25 |
| TCN1 | 3.57617649 | 5.64E-25 |
| AQP3 | 1.33552128 | 9.80E-25 |
| SCEL | -1.3445939 | 2.70E-24 |
| RAB3B | -1.742531 | 3.20E-24 |
| RRM2 | 1.43028087 | 3.57E-24 |
| F3 | -1.4835759 | 4.30E-24 |
| OASL | 2.39426404 | 5.06E-24 |
| DNASE1L3 | 1.22474701 | 5.50E-24 |
| MMP12 | 3.66631191 | 8.07E-24 |
| C14orf132 | -1.2942752 | 8.43E-24 |
| LTF | 3.50329731 | 1.21E-23 |
| CYP39A1 | -1.147887 | 1.39E-23 |
| PI3 | 3.41117684 | 2.05E-23 |
| PSMB10 | 1.06846557 | 2.05E-23 |
| FHL1 | -1.1861615 | 2.05E-23 |
| IFITM1 | 1.27361385 | 2.42E-23 |
| AQP9 | -2.077693 | 3.36E-23 |
| TNFSF10 | 1.00139729 | 3.51E-23 |
| FUT3 | 1.54605081 | 5.34E-23 |
| PRSS22 | 1.5905916 | 6.30E-23 |
| ID4 | -1.2806382 | 6.73E-23 |
| ZBED2 | 1.73492125 | 6.82E-23 |
| SPINK1 | -1.6632268 | 6.86E-23 |
| CYP2J2 | -1.5908564 | 8.31E-23 |
| TXNIP | -1.0588111 | 9.78E-23 |
| MATK | 1.39506302 | 9.78E-23 |
| OAS1 | 1.48161319 | 1.08E-22 |
| WIF1 | -2.4922676 | 1.32E-22 |
| TNFRSF21 | 1.49156704 | 1.41E-22 |
| CLEC7A | 1.38654443 | 2.71E-22 |
| MMP1 | 3.0574964 | 3.65E-22 |
| MX1 | 1.78796732 | 4.02E-22 |
| PELI2 | -1.1208595 | 4.32E-22 |
| SPRR1B | 2.55772097 | 4.59E-22 |
| F12 | 1.0098174 | 7.02E-22 |
| FUT2 | 1.05313134 | 9.77E-22 |
| ACADL | -1.2525486 | 1.02E-21 |
| NUAK1 | -1.0696888 | 1.09E-21 |
| NELL2 | 2.43801977 | 1.28E-21 |
| RGS20 | 1.44540747 | 1.38E-21 |
| CDH3 | 1.37046967 | 1.38E-21 |
| SELE | 1.89474083 | 1.54E-21 |
| UPP1 | 1.40540673 | 2.00E-21 |
| FSCN1 | 1.04413603 | 2.02E-21 |
| CCL13 | 1.92427516 | 2.02E-21 |
| SERPINB13 | 1.38815935 | 2.48E-21 |
| IL20RA | -1.1533193 | 3.25E-21 |
| MSMB | -2.2320249 | 5.39E-21 |
| CDH12 | -1.7252473 | 5.70E-21 |
| OAS2 | 1.59752754 | 6.16E-21 |
| ZNF135 | -1.1022188 | 7.07E-21 |
| COL4A4 | 1.19044037 | 1.09E-20 |
| PLA2G3 | 1.31078885 | 1.82E-20 |
| SOCS3 | 1.25499731 | 1.85E-20 |
| NOD2 | 1.03598422 | 2.15E-20 |
| IRF9 | 1.01718171 | 2.16E-20 |
| NR3C2 | -1.0763081 | 2.30E-20 |
| CLEC10A | 1.22815668 | 2.36E-20 |
| ADAM19 | 1.09357115 | 2.36E-20 |
| CHP2 | -1.3600886 | 2.80E-20 |
| INA | 1.24951296 | 2.94E-20 |
| SPRR1A | 2.53453322 | 5.70E-20 |
| PAK3 | -1.0905447 | 6.11E-20 |
| HERC6 | 1.46846019 | 6.79E-20 |
| TNNC1 | -1.6244167 | 1.29E-19 |
| FOXM1 | 1.03067706 | 1.48E-19 |
| KRT15 | -1.4987875 | 1.55E-19 |
| SPRR2C | 2.52672294 | 1.98E-19 |
| ANGPTL4 | 1.58635863 | 1.98E-19 |
| APOE | -1.2636853 | 2.01E-19 |
| CLDN8 | -1.6092454 | 2.40E-19 |
| IFITM3 | 1.02178761 | 2.65E-19 |
| ARNTL2 | 1.23408603 | 3.03E-19 |
| CHI3L2 | 2.15097371 | 3.13E-19 |
| SNTB1 | -1.203939 | 4.31E-19 |
| OAS3 | 1.17279674 | 4.35E-19 |
| CD1B | 2.00086465 | 5.41E-19 |
| FABP5 | 1.52424533 | 5.84E-19 |
| CCR7 | 1.7327244 | 5.87E-19 |
| KRT6B | 1.98631943 | 6.12E-19 |
| CCNB1 | 1.14648338 | 6.36E-19 |
| ELF3 | 1.06857779 | 6.99E-19 |
| IFI6 | 1.82707334 | 7.02E-19 |
| TGM3 | 1.25391117 | 7.15E-19 |
| KYNU | 1.34054927 | 1.14E-18 |
| FHOD3 | -1.0948231 | 1.41E-18 |
| CEP55 | 1.1336864 | 1.79E-18 |
| TRIM15 | 1.52671754 | 2.55E-18 |
| FOSL1 | 1.70824196 | 2.86E-18 |
| MMP9 | 1.43897238 | 4.09E-18 |
| CKS2 | 1.02160896 | 5.21E-18 |
| TGM1 | 1.17075346 | 1.36E-17 |
| CYP2W1 | -1.2419081 | 2.11E-17 |
| HS3ST3A1 | 1.73394168 | 2.23E-17 |
| RGS1 | 1.66686091 | 2.38E-17 |
| FABP7 | -2.360763 | 2.74E-17 |
| SLAMF1 | 1.07947373 | 2.88E-17 |
| GSTM3 | -1.0218381 | 3.59E-17 |
| GDPD3 | 1.14317 | 5.25E-17 |
| IL13 | 1.70560695 | 5.83E-17 |
| SAMSN1 | 1.32571603 | 6.58E-17 |
| NPY1R | -1.1841455 | 7.44E-17 |
| ITGAX | 1.0227722 | 7.83E-17 |
| APOL1 | 1.12281101 | 8.05E-17 |
| ISG15 | 1.61985851 | 9.16E-17 |
| SLAMF7 | 1.02375843 | 1.25E-16 |
| APOBEC3B | 1.38934634 | 1.27E-16 |
| RTP4 | 1.5471204 | 1.46E-16 |
| FCGBP | -1.0529591 | 1.70E-16 |
| THY1 | 1.10921356 | 1.73E-16 |
| CA2 | 1.4269749 | 2.58E-16 |
| CD1C | 1.18495299 | 2.59E-16 |
| SCIN | -1.1947451 | 3.57E-16 |
| TRIM10 | 1.12239397 | 4.21E-16 |
| MATN4 | -1.1347812 | 4.34E-16 |
| ALOX15 | 1.24876763 | 6.20E-16 |
| ARL14 | 1.55566964 | 8.32E-16 |
| SRGN | 1.0121285 | 8.61E-16 |
| CCL22 | 1.68266968 | 9.47E-16 |
| S100A12 | 2.39052984 | 9.86E-16 |
| CACNA1H | -1.6430032 | 1.07E-15 |
| GZMB | 1.66926603 | 1.28E-15 |
| LOR | -1.1409224 | 1.51E-15 |
| CH25H | 1.62053074 | 1.64E-15 |
| KCNK10 | 1.1263879 | 1.71E-15 |
| CXCR4 | 1.14380045 | 2.00E-15 |
| CCR5 | 1.10473627 | 2.02E-15 |
| IL32 | 1.17454112 | 2.05E-15 |
| GAN | -1.0616033 | 2.05E-15 |
| CA3 | -1.0232772 | 2.70E-15 |
| CST7 | 1.28203095 | 4.09E-15 |
| KREMEN2 | 1.06912822 | 4.17E-15 |
| IFI44 | 1.36277292 | 4.32E-15 |
| PPP1R3C | 1.40664786 | 4.46E-15 |
| CXCL10 | 2.10622954 | 4.66E-15 |
| PON3 | -1.1406556 | 4.66E-15 |
| IL13RA2 | 1.5489117 | 5.27E-15 |
| CD3D | 1.23963913 | 7.24E-15 |
| ADAMDEC1 | 1.5995163 | 7.24E-15 |
| SLC5A1 | 1.01794145 | 9.41E-15 |
| CXCL1 | 2.3253611 | 1.02E-14 |
| BIRC3 | 1.18341133 | 1.03E-14 |
| TNFRSF12A | 1.24866106 | 1.04E-14 |
| GGH | 1.14128157 | 1.11E-14 |
| CCL19 | 1.11891982 | 1.11E-14 |
| IL7R | 1.19640235 | 1.38E-14 |
| TSPAN8 | -1.335816 | 1.46E-14 |
| LCN2 | 1.86253771 | 1.48E-14 |
| CCL2 | 1.55964584 | 3.14E-14 |
| PLEK | 1.00316499 | 3.29E-14 |
| BATF3 | 1.30113281 | 3.39E-14 |
| CYP27B1 | 1.01813574 | 3.90E-14 |
| APOD | -1.0663451 | 3.91E-14 |
| OGN | -1.2837808 | 4.02E-14 |
| NETO2 | 1.15725453 | 5.19E-14 |
| AIM2 | 1.13806972 | 7.03E-14 |
| PRSS3 | 1.1119864 | 7.49E-14 |
| HTR3A | 1.4129879 | 7.71E-14 |
| NTRK1 | 1.47425204 | 1.41E-13 |
| PITX1 | 1.18804508 | 1.59E-13 |
| HPSE | 1.15533862 | 2.00E-13 |
| MMP3 | 1.97729742 | 2.96E-13 |
| RHCG | 1.57292585 | 3.42E-13 |
| HBEGF | 1.00336015 | 3.72E-13 |
| ADAM8 | 1.02944022 | 4.36E-13 |
| TMC5 | 1.13161335 | 4.84E-13 |
| XAF1 | 1.13421275 | 6.10E-13 |
| CD3G | 1.02711215 | 6.21E-13 |
| MDFI | 1.01687571 | 6.71E-13 |
| PNPLA3 | -1.194514 | 7.39E-13 |
| IFIT3 | 1.00986792 | 8.20E-13 |
| ITK | 1.02382403 | 8.97E-13 |
| CHRNA3 | 1.03926996 | 1.49E-12 |
| CD2 | 1.13569538 | 1.62E-12 |
| COMP | 1.49952617 | 4.28E-12 |
| IL2RA | 1.06376398 | 8.16E-12 |
| CXCL9 | 1.8410822 | 1.04E-11 |
| TMPRSS11D | 1.40893815 | 1.21E-11 |
| IL26 | 1.22543788 | 1.40E-11 |
| CCL17 | 1.52243029 | 1.41E-11 |
| PSORS1C2 | -1.0336868 | 1.43E-11 |
| CXCL11 | 1.45627001 | 1.47E-11 |
| SNX10 | 1.03921689 | 1.93E-11 |
| ERBB4 | -1.0357658 | 2.42E-11 |
| LYZ | 1.31145538 | 2.42E-11 |
| ZBTB16 | -1.264694 | 2.81E-11 |
| HAL | 1.09562674 | 3.02E-11 |
| PDK4 | -1.0134088 | 3.34E-11 |
| GPR171 | 1.04270861 | 3.40E-11 |
| ELF5 | 1.13621079 | 8.63E-11 |
| ATP12A | 1.46569793 | 8.84E-11 |
| HMGCS2 | -1.1100864 | 1.07E-10 |
| KLK6 | 1.65384957 | 1.43E-10 |
| CD69 | 1.11966858 | 3.90E-10 |
| CHRM4 | -2.4572997 | 5.69E-10 |
| IFI44L | 1.16815868 | 1.10E-09 |
| SLC5A5 | 1.23185155 | 1.39E-09 |
| IL22 | 1.12032191 | 2.07E-09 |
| HBB | -1.1694898 | 2.39E-09 |
| CD177 | 1.22882865 | 2.63E-09 |
| TNNI2 | -1.4343388 | 4.31E-09 |
| CHRNA9 | 1.01331845 | 8.19E-09 |
| CAMP | 1.01845163 | 8.98E-09 |
| GZMA | 1.11641269 | 9.54E-09 |
| FMO2 | 1.0999154 | 9.54E-09 |
| CHAC1 | 1.07819898 | 9.80E-09 |
| RARRES1 | -1.1300138 | 1.21E-08 |
| RSAD2 | 1.14150589 | 2.18E-08 |
| PDZK1 | -1.4090361 | 2.74E-08 |
| PLA2G2A | 1.24876882 | 3.32E-08 |
| S100P | 1.04786638 | 3.93E-08 |
| FADS2 | -1.6109721 | 1.33E-07 |
| SPRR3 | 1.0207871 | 2.34E-07 |
| APOC1 | -1.3290755 | 2.36E-07 |
| PSG4 | -1.0478514 | 4.85E-07 |
| CCL20 | 1.03060152 | 7.00E-07 |
| BMP3 | -1.9016523 | 8.89E-07 |
| ABCB11 | -1.7262685 | 1.69E-06 |
| ADH6 | -1.9095621 | 3.74E-06 |
| GAL | -1.7579176 | 5.18E-06 |
| HAND2 | -1.4635847 | 7.76E-06 |
| GJB4 | -2.0270868 | 3.47E-05 |
| HAO2 | -1.0749767 | 8.94E-05 |
| CLDN16 | -1.3634711 | 0.00011409 |
| XIST | -2.3266234 | 0.00015869 |
| AMOT | -1.9205428 | 0.00018118 |
| MAST1 | -1.3767947 | 0.00020961 |
| P2RY4 | -1.2430038 | 0.00023511 |
| SEMA3B | -1.8013092 | 0.00036125 |
| TLL1 | -1.4958571 | 0.00049325 |
| CDH20 | -1.2547594 | 0.00051296 |
| PRKY | 1.24560275 | 0.00100223 |
| TAS2R4 | -1.0814847 | 0.00121 |
| GLRA3 | -1.208374 | 0.00140199 |
| GALNT8 | -1.2440375 | 0.00140457 |
| LRP2BP | -1.3139286 | 0.00190128 |
| MASP2 | -1.1167011 | 0.00261865 |
| GSG1 | -1.0736659 | 0.00301473 |
| PHLDB1 | -1.7431881 | 0.00323919 |
| GAL3ST1 | -1.1987199 | 0.00415179 |
| ZNF549 | -1.6592288 | 0.00473172 |
| NCAM2 | -1.2306278 | 0.00556827 |
| ALOX12P2 | -1.1067348 | 0.00582812 |
| ALAS2 | -1.3113273 | 0.00744851 |
| SH3TC2 | -1.0030639 | 0.00874954 |
| RNF5 | -1.555698 | 0.01131346 |
| MTTP | -1.1454287 | 0.01747274 |
| MED24 | -1.3628939 | 0.03001847 |
| RPS4Y1 | 1.45100845 | 0.03501945 |
| CDKL5 | -1.0073697 | 0.04613798 |
| SOX21 | -1.2244902 | 0.04631141 |

**Table S3.** GO analysis of differentially expressed genes

| Ontology | ID | Description | qvalue |
| --- | --- | --- | --- |
| BP | GO:0019221 | cytokine-mediated signaling pathway | 4.81E-11 |
| BP | GO:0009615 | response to virus | 4.81E-11 |
| BP | GO:0097530 | granulocyte migration | 6.30E-11 |
| BP | GO:1990266 | neutrophil migration | 1.85E-10 |
| BP | GO:0071621 | granulocyte chemotaxis | 2.28E-10 |
| BP | GO:0030595 | leukocyte chemotaxis | 2.76E-10 |
| BP | GO:0051607 | defense response to virus | 4.94E-10 |
| BP | GO:0140546 | defense response to symbiont | 4.94E-10 |
| BP | GO:0050900 | leukocyte migration | 4.94E-10 |
| BP | GO:0030593 | neutrophil chemotaxis | 7.11E-10 |
| BP | GO:0048247 | lymphocyte chemotaxis | 2.17E-09 |
| BP | GO:0070098 | chemokine-mediated signaling pathway | 9.52E-09 |
| BP | GO:0060326 | cell chemotaxis | 9.52E-09 |
| BP | GO:0071674 | mononuclear cell migration | 2.12E-08 |
| BP | GO:0097529 | myeloid leukocyte migration | 2.12E-08 |
| BP | GO:1990868 | response to chemokine | 2.77E-08 |
| BP | GO:1990869 | cellular response to chemokine | 2.77E-08 |
| BP | GO:0072676 | lymphocyte migration | 3.24E-08 |
| BP | GO:0019730 | antimicrobial humoral response | 5.60E-08 |
| BP | GO:0045071 | negative regulation of viral genome replication | 7.26E-08 |
| BP | GO:0034340 | response to type I interferon | 1.03E-07 |
| BP | GO:0048525 | negative regulation of viral process | 1.28E-07 |
| BP | GO:0061844 | antimicrobial humoral immune response mediated by antimicrobial peptide | 2.31E-07 |
| BP | GO:0019058 | viral life cycle | 2.69E-07 |
| BP | GO:0060337 | type I interferon signaling pathway | 3.00E-07 |
| BP | GO:0050792 | regulation of viral process | 3.29E-07 |
| BP | GO:0001906 | cell killing | 3.29E-07 |
| BP | GO:0071357 | cellular response to type I interferon | 4.01E-07 |
| BP | GO:0070374 | positive regulation of ERK1 and ERK2 cascade | 4.16E-07 |
| BP | GO:1903900 | regulation of viral life cycle | 5.37E-07 |
| BP | GO:0030216 | keratinocyte differentiation | 1.83E-06 |
| BP | GO:0045069 | regulation of viral genome replication | 4.44E-06 |
| BP | GO:0006959 | humoral immune response | 5.22E-06 |
| BP | GO:0016032 | viral process | 5.53E-06 |
| BP | GO:0019079 | viral genome replication | 6.08E-06 |
| BP | GO:0002548 | monocyte chemotaxis | 6.08E-06 |
| BP | GO:0050727 | regulation of inflammatory response | 6.08E-06 |
| BP | GO:0043588 | skin development | 6.25E-06 |
| BP | GO:0032103 | positive regulation of response to external stimulus | 8.06E-06 |
| BP | GO:0001819 | positive regulation of cytokine production | 9.86E-06 |
| BP | GO:0070372 | regulation of ERK1 and ERK2 cascade | 1.35E-05 |
| BP | GO:0043410 | positive regulation of MAPK cascade | 1.55E-05 |
| BP | GO:0008544 | epidermis development | 2.67E-05 |
| BP | GO:0098586 | cellular response to virus | 2.94E-05 |
| BP | GO:0070371 | ERK1 and ERK2 cascade | 3.37E-05 |
| BP | GO:0034341 | response to interferon-gamma | 7.96E-05 |
| BP | GO:0009913 | epidermal cell differentiation | 0.00012189 |
| BP | GO:0071887 | leukocyte apoptotic process | 0.00022524 |
| BP | GO:0032496 | response to lipopolysaccharide | 0.00022524 |
| BP | GO:0051651 | maintenance of location in cell | 0.00022524 |
| BP | GO:0045088 | regulation of innate immune response | 0.00027368 |
| BP | GO:0042742 | defense response to bacterium | 0.00028652 |
| BP | GO:0042060 | wound healing | 0.00028764 |
| BP | GO:0022409 | positive regulation of cell-cell adhesion | 0.00029394 |
| BP | GO:0072678 | T cell migration | 0.00029394 |
| BP | GO:0002274 | myeloid leukocyte activation | 0.00032365 |
| BP | GO:0019835 | cytolysis | 0.00032399 |
| BP | GO:0002523 | leukocyte migration involved in inflammatory response | 0.00032681 |
| BP | GO:0031640 | killing of cells of other organism | 0.00034373 |
| BP | GO:0002703 | regulation of leukocyte mediated immunity | 0.00035206 |
| BP | GO:0002683 | negative regulation of immune system process | 0.00037817 |
| BP | GO:0002831 | regulation of response to biotic stimulus | 0.00038185 |
| BP | GO:0002237 | response to molecule of bacterial origin | 0.00038996 |
| BP | GO:0018149 | peptide cross-linking | 0.00049051 |
| BP | GO:0071605 | monocyte chemotactic protein-1 production | 0.00049051 |
| BP | GO:0071637 | regulation of monocyte chemotactic protein-1 production | 0.00049051 |
| BP | GO:0042110 | T cell activation | 0.00055537 |
| BP | GO:0060338 | regulation of type I interferon-mediated signaling pathway | 0.00055537 |
| BP | GO:0032069 | regulation of nuclease activity | 0.00059984 |
| BP | GO:0002685 | regulation of leukocyte migration | 0.00064351 |
| BP | GO:0051092 | positive regulation of NF-kappaB transcription factor activity | 0.0006759 |
| BP | GO:0050867 | positive regulation of cell activation | 0.0006995 |
| BP | GO:0031349 | positive regulation of defense response | 0.00071559 |
| BP | GO:0002819 | regulation of adaptive immune response | 0.00072883 |
| BP | GO:0009410 | response to xenobiotic stimulus | 0.00076093 |
| BP | GO:0031424 | keratinization | 0.00083757 |
| BP | GO:0032481 | positive regulation of type I interferon production | 0.00083757 |
| BP | GO:2000106 | regulation of leukocyte apoptotic process | 0.00095793 |
| BP | GO:0043547 | positive regulation of GTPase activity | 0.00103691 |
| BP | GO:0044403 | biological process involved in symbiotic interaction | 0.00106411 |
| BP | GO:1903557 | positive regulation of tumor necrosis factor superfamily cytokine production | 0.00106411 |
| BP | GO:2000401 | regulation of lymphocyte migration | 0.00109873 |
| BP | GO:0051235 | maintenance of location | 0.00110183 |
| BP | GO:2000404 | regulation of T cell migration | 0.00111742 |
| BP | GO:0002443 | leukocyte mediated immunity | 0.00111742 |
| BP | GO:0050777 | negative regulation of immune response | 0.00111742 |
| BP | GO:0002407 | dendritic cell chemotaxis | 0.00113193 |
| BP | GO:0002696 | positive regulation of leukocyte activation | 0.00136401 |
| BP | GO:0071347 | cellular response to interleukin-1 | 0.00149254 |
| BP | GO:0071677 | positive regulation of mononuclear cell migration | 0.00151879 |
| BP | GO:0002697 | regulation of immune effector process | 0.00154692 |
| BP | GO:0002699 | positive regulation of immune effector process | 0.00158486 |
| BP | GO:0071675 | regulation of mononuclear cell migration | 0.00163964 |
| BP | GO:0070555 | response to interleukin-1 | 0.00165 |
| BP | GO:2000406 | positive regulation of T cell migration | 0.00180449 |
| BP | GO:0043270 | positive regulation of ion transport | 0.00193829 |
| BP | GO:0034368 | protein-lipid complex remodeling | 0.00206763 |
| BP | GO:0034369 | plasma lipoprotein particle remodeling | 0.00206763 |
| BP | GO:0034374 | low-density lipoprotein particle remodeling | 0.00206763 |
| BP | GO:0051238 | sequestering of metal ion | 0.00206763 |
| BP | GO:0071639 | positive regulation of monocyte chemotactic protein-1 production | 0.00206763 |
| BP | GO:0071216 | cellular response to biotic stimulus | 0.00227711 |
| BP | GO:0043903 | regulation of biological process involved in symbiotic interaction | 0.00257072 |
| BP | GO:0051797 | regulation of hair follicle development | 0.00259524 |
| BP | GO:0034367 | protein-containing complex remodeling | 0.00264437 |
| BP | GO:0035456 | response to interferon-beta | 0.00264437 |
| BP | GO:0071706 | tumor necrosis factor superfamily cytokine production | 0.00279921 |
| BP | GO:1903555 | regulation of tumor necrosis factor superfamily cytokine production | 0.00279921 |
| BP | GO:0034612 | response to tumor necrosis factor | 0.00279921 |
| BP | GO:0097305 | response to alcohol | 0.00279921 |
| BP | GO:0031341 | regulation of cell killing | 0.00284722 |
| BP | GO:0036336 | dendritic cell migration | 0.00291408 |
| BP | GO:0039528 | cytoplasmic pattern recognition receptor signaling pathway in response to virus | 0.00334548 |
| BP | GO:0007159 | leukocyte cell-cell adhesion | 0.00353452 |
| BP | GO:0032760 | positive regulation of tumor necrosis factor production | 0.00362692 |
| BP | GO:0010038 | response to metal ion | 0.00369347 |
| BP | GO:2000403 | positive regulation of lymphocyte migration | 0.00372423 |
| BP | GO:0071356 | cellular response to tumor necrosis factor | 0.00397889 |
| BP | GO:0002886 | regulation of myeloid leukocyte mediated immunity | 0.00412651 |
| BP | GO:0061041 | regulation of wound healing | 0.00412803 |
| BP | GO:0002687 | positive regulation of leukocyte migration | 0.00427818 |
| BP | GO:0003159 | morphogenesis of an endothelium | 0.00427818 |
| BP | GO:0060339 | negative regulation of type I interferon-mediated signaling pathway | 0.00427818 |
| BP | GO:0061154 | endothelial tube morphogenesis | 0.00427818 |
| BP | GO:0032102 | negative regulation of response to external stimulus | 0.00428148 |
| BP | GO:0052547 | regulation of peptidase activity | 0.00430567 |
| BP | GO:0034767 | positive regulation of ion transmembrane transport | 0.0044267 |
| BP | GO:0002833 | positive regulation of response to biotic stimulus | 0.00461006 |
| BP | GO:1901617 | organic hydroxy compound biosynthetic process | 0.00497424 |
| BP | GO:0043087 | regulation of GTPase activity | 0.00497424 |
| BP | GO:0002820 | negative regulation of adaptive immune response | 0.00497424 |
| BP | GO:0090303 | positive regulation of wound healing | 0.00497424 |
| BP | GO:1903039 | positive regulation of leukocyte cell-cell adhesion | 0.00523856 |
| BP | GO:0002753 | cytoplasmic pattern recognition receptor signaling pathway | 0.00537747 |
| BP | GO:0032728 | positive regulation of interferon-beta production | 0.00543052 |
| BP | GO:0002526 | acute inflammatory response | 0.00543052 |
| BP | GO:0052548 | regulation of endopeptidase activity | 0.00543052 |
| BP | GO:0050729 | positive regulation of inflammatory response | 0.00554528 |
| BP | GO:0019748 | secondary metabolic process | 0.00567885 |
| BP | GO:0052126 | movement in host environment | 0.00585593 |
| BP | GO:0051281 | positive regulation of release of sequestered calcium ion into cytosol | 0.00589608 |
| BP | GO:0071222 | cellular response to lipopolysaccharide | 0.00591942 |
| BP | GO:0030278 | regulation of ossification | 0.00618481 |
| BP | GO:0051209 | release of sequestered calcium ion into cytosol | 0.00618481 |
| BP | GO:0071466 | cellular response to xenobiotic stimulus | 0.00619142 |
| BP | GO:0022617 | extracellular matrix disassembly | 0.00642514 |
| BP | GO:0051283 | negative regulation of sequestering of calcium ion | 0.00642514 |
| BP | GO:0044848 | biological phase | 0.00674899 |
| BP | GO:0030574 | collagen catabolic process | 0.00700815 |
| BP | GO:0051282 | regulation of sequestering of calcium ion | 0.00700815 |
| BP | GO:0071346 | cellular response to interferon-gamma | 0.00700815 |
| BP | GO:0032640 | tumor necrosis factor production | 0.00700815 |
| BP | GO:0032680 | regulation of tumor necrosis factor production | 0.00700815 |
| BP | GO:0043302 | positive regulation of leukocyte degranulation | 0.00717705 |
| BP | GO:0050870 | positive regulation of T cell activation | 0.00717705 |
| BP | GO:0022407 | regulation of cell-cell adhesion | 0.00724553 |
| BP | GO:0002275 | myeloid cell activation involved in immune response | 0.00729598 |
| BP | GO:0001503 | ossification | 0.00738007 |
| BP | GO:0044409 | entry into host | 0.00752584 |
| BP | GO:1904064 | positive regulation of cation transmembrane transport | 0.00752584 |
| BP | GO:0034764 | positive regulation of transmembrane transport | 0.00776228 |
| BP | GO:0019216 | regulation of lipid metabolic process | 0.00780365 |
| BP | GO:1901623 | regulation of lymphocyte chemotaxis | 0.00794364 |
| BP | GO:1903901 | negative regulation of viral life cycle | 0.00794364 |
| BP | GO:0002688 | regulation of leukocyte chemotaxis | 0.00794364 |
| BP | GO:0051208 | sequestering of calcium ion | 0.00794364 |
| BP | GO:0030217 | T cell differentiation | 0.00794364 |
| BP | GO:0071219 | cellular response to molecule of bacterial origin | 0.00794364 |
| BP | GO:0002604 | regulation of dendritic cell antigen processing and presentation | 0.00794364 |
| BP | GO:0051798 | positive regulation of hair follicle development | 0.00794364 |
| BP | GO:0071492 | cellular response to UV-A | 0.00794364 |
| BP | GO:1900046 | regulation of hemostasis | 0.00829668 |
| BP | GO:0050920 | regulation of chemotaxis | 0.00840819 |
| BP | GO:0001909 | leukocyte mediated cytotoxicity | 0.00841327 |
| BP | GO:0032479 | regulation of type I interferon production | 0.00841327 |
| BP | GO:0032606 | type I interferon production | 0.00841327 |
| BP | GO:0051091 | positive regulation of DNA-binding transcription factor activity | 0.00841327 |
| BP | GO:0006706 | steroid catabolic process | 0.00852599 |
| BP | GO:0042634 | regulation of hair cycle | 0.00852599 |
| BP | GO:0016485 | protein processing | 0.00870641 |
| BP | GO:0051346 | negative regulation of hydrolase activity | 0.00875421 |
| BP | GO:0043300 | regulation of leukocyte degranulation | 0.00875421 |
| BP | GO:0071827 | plasma lipoprotein particle organization | 0.00875421 |
| BP | GO:2000107 | negative regulation of leukocyte apoptotic process | 0.00875421 |
| BP | GO:0032874 | positive regulation of stress-activated MAPK cascade | 0.00888846 |
| BP | GO:0030099 | myeloid cell differentiation | 0.00906631 |
| BP | GO:0032872 | regulation of stress-activated MAPK cascade | 0.00910768 |
| BP | GO:0039529 | RIG-I signaling pathway | 0.00941917 |
| BP | GO:0032642 | regulation of chemokine production | 0.00944852 |
| BP | GO:0032722 | positive regulation of chemokine production | 0.00944852 |
| BP | GO:0045229 | external encapsulating structure organization | 0.00944852 |
| BP | GO:0071695 | anatomical structure maturation | 0.00944852 |
| BP | GO:0070304 | positive regulation of stress-activated protein kinase signaling cascade | 0.00945586 |
| BP | GO:1903131 | mononuclear cell differentiation | 0.00957903 |
| BP | GO:0032602 | chemokine production | 0.00977223 |
| BP | GO:0070302 | regulation of stress-activated protein kinase signaling cascade | 0.00979231 |
| BP | GO:0070227 | lymphocyte apoptotic process | 0.00979231 |
| BP | GO:1903036 | positive regulation of response to wounding | 0.00979231 |
| BP | GO:0010818 | T cell chemotaxis | 0.01021002 |
| BP | GO:0031342 | negative regulation of cell killing | 0.01021002 |
| BP | GO:0045089 | positive regulation of innate immune response | 0.01057512 |
| BP | GO:0042304 | regulation of fatty acid biosynthetic process | 0.01059392 |
| BP | GO:0050832 | defense response to fungus | 0.01059392 |
| BP | GO:0052372 | modulation by symbiont of entry into host | 0.01059392 |
| BP | GO:0002449 | lymphocyte mediated immunity | 0.01066014 |
| BP | GO:0046470 | phosphatidylcholine metabolic process | 0.01084426 |
| BP | GO:2000116 | regulation of cysteine-type endopeptidase activity | 0.01084426 |
| BP | GO:0015791 | polyol transport | 0.01091568 |
| BP | GO:0019755 | one-carbon compound transport | 0.01091568 |
| BP | GO:0048820 | hair follicle maturation | 0.01091568 |
| BP | GO:0060340 | positive regulation of type I interferon-mediated signaling pathway | 0.01091568 |
| BP | GO:0071825 | protein-lipid complex subunit organization | 0.01119114 |
| BP | GO:0045785 | positive regulation of cell adhesion | 0.01143057 |
| BP | GO:0042552 | myelination | 0.01151789 |
| BP | GO:1903034 | regulation of response to wounding | 0.01156571 |
| BP | GO:0002822 | regulation of adaptive immune response based on somatic recombination of immune receptors built from immunoglobulin superfamily domains | 0.01192158 |
| BP | GO:0071622 | regulation of granulocyte chemotaxis | 0.01192158 |
| BP | GO:1903307 | positive regulation of regulated secretory pathway | 0.01192158 |
| BP | GO:0051701 | biological process involved in interaction with host | 0.01193586 |
| BP | GO:0034694 | response to prostaglandin | 0.01202806 |
| BP | GO:0043032 | positive regulation of macrophage activation | 0.01202806 |
| BP | GO:0043304 | regulation of mast cell degranulation | 0.01202806 |
| BP | GO:0062207 | regulation of pattern recognition receptor signaling pathway | 0.01206293 |
| BP | GO:0007272 | ensheathment of neurons | 0.01206293 |
| BP | GO:0008366 | axon ensheathment | 0.01206293 |
| BP | GO:0033008 | positive regulation of mast cell activation involved in immune response | 0.01266881 |
| BP | GO:0043306 | positive regulation of mast cell degranulation | 0.01266881 |
| BP | GO:0070141 | response to UV-A | 0.01266881 |
| BP | GO:0016042 | lipid catabolic process | 0.01266881 |
| BP | GO:0042303 | molting cycle | 0.01300581 |
| BP | GO:0042633 | hair cycle | 0.01300581 |
| BP | GO:0033006 | regulation of mast cell activation involved in immune response | 0.01300581 |
| BP | GO:0070633 | transepithelial transport | 0.01300581 |
| BP | GO:0002221 | pattern recognition receptor signaling pathway | 0.01308411 |
| BP | GO:1901184 | regulation of ERBB signaling pathway | 0.01346511 |
| BP | GO:0002456 | T cell mediated immunity | 0.0142395 |
| BP | GO:0010524 | positive regulation of calcium ion transport into cytosol | 0.0142395 |
| BP | GO:0050921 | positive regulation of chemotaxis | 0.01438952 |
| BP | GO:1903706 | regulation of hemopoiesis | 0.01438952 |
| BP | GO:0002468 | dendritic cell antigen processing and presentation | 0.01481587 |
| BP | GO:0140374 | antiviral innate immune response | 0.01481587 |
| BP | GO:0097553 | calcium ion transmembrane import into cytosol | 0.01481587 |
| BP | GO:0001942 | hair follicle development | 0.0148341 |
| BP | GO:0050863 | regulation of T cell activation | 0.01526462 |
| BP | GO:0006805 | xenobiotic metabolic process | 0.01526885 |
| BP | GO:0046718 | viral entry into host cell | 0.01585044 |
| BP | GO:0032609 | interferon-gamma production | 0.01585044 |
| BP | GO:0032649 | regulation of interferon-gamma production | 0.01585044 |
| BP | GO:0010951 | negative regulation of endopeptidase activity | 0.01585044 |
| BP | GO:0032608 | interferon-beta production | 0.01585378 |
| BP | GO:0032648 | regulation of interferon-beta production | 0.01585378 |
| BP | GO:0030098 | lymphocyte differentiation | 0.01629873 |
| BP | GO:0055074 | calcium ion homeostasis | 0.01637816 |
| BP | GO:0009612 | response to mechanical stimulus | 0.01641807 |
| BP | GO:0022404 | molting cycle process | 0.01671836 |
| BP | GO:0022405 | hair cycle process | 0.01671836 |
| BP | GO:0002830 | positive regulation of type 2 immune response | 0.01671836 |
| BP | GO:0010819 | regulation of T cell chemotaxis | 0.01671836 |
| BP | GO:0034375 | high-density lipoprotein particle remodeling | 0.01671836 |
| BP | GO:0051044 | positive regulation of membrane protein ectodomain proteolysis | 0.01671836 |
| BP | GO:1902931 | negative regulation of alcohol biosynthetic process | 0.01671836 |
| BP | GO:1903037 | regulation of leukocyte cell-cell adhesion | 0.01714531 |
| BP | GO:0002709 | regulation of T cell mediated immunity | 0.01745093 |
| BP | GO:0097006 | regulation of plasma lipoprotein particle levels | 0.01745093 |
| BP | GO:0098773 | skin epidermis development | 0.01745093 |
| BP | GO:0031348 | negative regulation of defense response | 0.01787795 |
| BP | GO:0033280 | response to vitamin D | 0.01802878 |
| BP | GO:0045921 | positive regulation of exocytosis | 0.01832315 |
| BP | GO:0002764 | immune response-regulating signaling pathway | 0.01835209 |
| BP | GO:0002702 | positive regulation of production of molecular mediator of immune response | 0.01876993 |
| BP | GO:0043123 | positive regulation of I-kappaB kinase/NF-kappaB signaling | 0.01942037 |
| BP | GO:0045742 | positive regulation of epidermal growth factor receptor signaling pathway | 0.01965709 |
| BP | GO:0010466 | negative regulation of peptidase activity | 0.01965709 |
| BP | GO:0009620 | response to fungus | 0.01979041 |
| BP | GO:0030282 | bone mineralization | 0.02030186 |
| BP | GO:0034113 | heterotypic cell-cell adhesion | 0.02106227 |
| BP | GO:0043030 | regulation of macrophage activation | 0.02106227 |
| BP | GO:0051897 | positive regulation of protein kinase B signaling | 0.02106227 |
| BP | GO:0042178 | xenobiotic catabolic process | 0.02126351 |
| BP | GO:0032613 | interleukin-10 production | 0.02193176 |
| BP | GO:0032615 | interleukin-12 production | 0.02193176 |
| BP | GO:0032653 | regulation of interleukin-10 production | 0.02193176 |
| BP | GO:0032655 | regulation of interleukin-12 production | 0.02193176 |
| BP | GO:0032731 | positive regulation of interleukin-1 beta production | 0.02193176 |
| BP | GO:0032757 | positive regulation of interleukin-8 production | 0.02193176 |
| BP | GO:0045576 | mast cell activation | 0.02193176 |
| BP | GO:0045582 | positive regulation of T cell differentiation | 0.02275704 |
| BP | GO:1901186 | positive regulation of ERBB signaling pathway | 0.02275704 |
| BP | GO:0045055 | regulated exocytosis | 0.02288295 |
| BP | GO:0051047 | positive regulation of secretion | 0.02304979 |
| BP | GO:0002704 | negative regulation of leukocyte mediated immunity | 0.02304979 |
| BP | GO:1902107 | positive regulation of leukocyte differentiation | 0.02304979 |
| BP | GO:1903708 | positive regulation of hemopoiesis | 0.02304979 |
| BP | GO:0072503 | cellular divalent inorganic cation homeostasis | 0.02447432 |
| BP | GO:0019217 | regulation of fatty acid metabolic process | 0.02451683 |
| BP | GO:0032695 | negative regulation of interleukin-12 production | 0.02451683 |
| BP | GO:0051546 | keratinocyte migration | 0.02451683 |
| BP | GO:1901739 | regulation of myoblast fusion | 0.02451683 |
| BP | GO:0060249 | anatomical structure homeostasis | 0.02479006 |
| BP | GO:0002460 | adaptive immune response based on somatic recombination of immune receptors built from immunoglobulin superfamily domains | 0.02479006 |
| BP | GO:0060402 | calcium ion transport into cytosol | 0.02522231 |
| BP | GO:0002690 | positive regulation of leukocyte chemotaxis | 0.0253233 |
| BP | GO:0051702 | biological process involved in interaction with symbiont | 0.0253233 |
| BP | GO:0002720 | positive regulation of cytokine production involved in immune response | 0.0253233 |
| BP | GO:0045453 | bone resorption | 0.0253233 |
| BP | GO:0150077 | regulation of neuroinflammatory response | 0.02591059 |
| BP | GO:0030193 | regulation of blood coagulation | 0.0268963 |
| BP | GO:0008202 | steroid metabolic process | 0.02716196 |
| BP | GO:0002577 | regulation of antigen processing and presentation | 0.02716196 |
| BP | GO:0033005 | positive regulation of mast cell activation | 0.02716196 |
| BP | GO:0039535 | regulation of RIG-I signaling pathway | 0.02716196 |
| BP | GO:0043651 | linoleic acid metabolic process | 0.02716196 |
| BP | GO:0071731 | response to nitric oxide | 0.02716196 |
| BP | GO:0051251 | positive regulation of lymphocyte activation | 0.0273208 |
| BP | GO:0032733 | positive regulation of interleukin-10 production | 0.02732428 |
| BP | GO:0032735 | positive regulation of interleukin-12 production | 0.02732428 |
| BP | GO:1902622 | regulation of neutrophil migration | 0.02732428 |
| BP | GO:0051403 | stress-activated MAPK cascade | 0.02732428 |
| BP | GO:1902105 | regulation of leukocyte differentiation | 0.02732428 |
| BP | GO:0002700 | regulation of production of molecular mediator of immune response | 0.02760527 |
| BP | GO:0021700 | developmental maturation | 0.02790199 |
| BP | GO:0120162 | positive regulation of cold-induced thermogenesis | 0.02800611 |
| BP | GO:0017157 | regulation of exocytosis | 0.02837105 |
| BP | GO:0007520 | myoblast fusion | 0.02910555 |
| BP | GO:0033003 | regulation of mast cell activation | 0.02910555 |
| BP | GO:0046596 | regulation of viral entry into host cell | 0.02910555 |
| BP | GO:0035455 | response to interferon-alpha | 0.02981974 |
| BP | GO:0046597 | negative regulation of viral entry into host cell | 0.02981974 |
| BP | GO:0070269 | pyroptosis | 0.02981974 |
| BP | GO:0002444 | myeloid leukocyte mediated immunity | 0.03023761 |
| BP | GO:0006509 | membrane protein ectodomain proteolysis | 0.03124255 |
| BP | GO:0019218 | regulation of steroid metabolic process | 0.03158486 |
| BP | GO:0045123 | cellular extravasation | 0.03201089 |
| BP | GO:0045862 | positive regulation of proteolysis | 0.03232575 |
| BP | GO:0002705 | positive regulation of leukocyte mediated immunity | 0.03239122 |
| BP | GO:0031098 | stress-activated protein kinase signaling cascade | 0.03267908 |
| BP | GO:0050830 | defense response to Gram-positive bacterium | 0.03267908 |
| BP | GO:0002573 | myeloid leukocyte differentiation | 0.03286358 |
| BP | GO:0045061 | thymic T cell selection | 0.03287917 |
| BP | GO:0045124 | regulation of bone resorption | 0.03287917 |
| BP | GO:0062208 | positive regulation of pattern recognition receptor signaling pathway | 0.03287917 |
| BP | GO:0150076 | neuroinflammatory response | 0.03287917 |
| BP | GO:0045824 | negative regulation of innate immune response | 0.03293854 |
| BP | GO:0050818 | regulation of coagulation | 0.03293854 |
| BP | GO:0046890 | regulation of lipid biosynthetic process | 0.03298845 |
| BP | GO:0018108 | peptidyl-tyrosine phosphorylation | 0.03331086 |
| BP | GO:0045833 | negative regulation of lipid metabolic process | 0.03331086 |
| BP | GO:0043122 | regulation of I-kappaB kinase/NF-kappaB signaling | 0.03344759 |
| BP | GO:1903305 | regulation of regulated secretory pathway | 0.03372222 |
| BP | GO:0044282 | small molecule catabolic process | 0.03372838 |
| BP | GO:0032729 | positive regulation of interferon-gamma production | 0.0342785 |
| BP | GO:0042180 | cellular ketone metabolic process | 0.03450569 |
| BP | GO:0043491 | protein kinase B signaling | 0.03450569 |
| BP | GO:0006816 | calcium ion transport | 0.03450569 |
| BP | GO:0045471 | response to ethanol | 0.03450569 |
| BP | GO:0018212 | peptidyl-tyrosine modification | 0.03465392 |
| BP | GO:0050878 | regulation of body fluid levels | 0.03473157 |
| BP | GO:0045621 | positive regulation of lymphocyte differentiation | 0.03473157 |
| BP | GO:0032732 | positive regulation of interleukin-1 production | 0.03473157 |
| BP | GO:0042058 | regulation of epidermal growth factor receptor signaling pathway | 0.03473157 |
| BP | GO:0043299 | leukocyte degranulation | 0.03473157 |
| BP | GO:1904427 | positive regulation of calcium ion transmembrane transport | 0.03473157 |
| BP | GO:0006833 | water transport | 0.03473157 |
| BP | GO:0030194 | positive regulation of blood coagulation | 0.03473157 |
| BP | GO:0034695 | response to prostaglandin E | 0.03473157 |
| BP | GO:0043931 | ossification involved in bone maturation | 0.03473157 |
| BP | GO:0072677 | eosinophil migration | 0.03473157 |
| BP | GO:1900048 | positive regulation of hemostasis | 0.03473157 |
| BP | GO:0051604 | protein maturation | 0.03499129 |
| BP | GO:0072330 | monocarboxylic acid biosynthetic process | 0.03611032 |
| BP | GO:0019882 | antigen processing and presentation | 0.03738534 |
| BP | GO:0035821 | modulation of process of other organism | 0.03738534 |
| BP | GO:0042116 | macrophage activation | 0.03738534 |
| BP | GO:0001667 | ameboidal-type cell migration | 0.03782325 |
| BP | GO:0031641 | regulation of myelination | 0.03782325 |
| BP | GO:0033077 | T cell differentiation in thymus | 0.03782325 |
| BP | GO:0034121 | regulation of toll-like receptor signaling pathway | 0.03782325 |
| BP | GO:0043627 | response to estrogen | 0.03782325 |
| BP | GO:0032928 | regulation of superoxide anion generation | 0.03782325 |
| BP | GO:0039531 | regulation of viral-induced cytoplasmic pattern recognition receptor signaling pathway | 0.03782325 |
| BP | GO:0050820 | positive regulation of coagulation | 0.03782325 |
| BP | GO:0051043 | regulation of membrane protein ectodomain proteolysis | 0.03782325 |
| BP | GO:0002824 | positive regulation of adaptive immune response based on somatic recombination of immune receptors built from immunoglobulin superfamily domains | 0.03800457 |
| BP | GO:0051341 | regulation of oxidoreductase activity | 0.03800457 |
| BP | GO:0002832 | negative regulation of response to biotic stimulus | 0.03946234 |
| BP | GO:0034440 | lipid oxidation | 0.03946234 |
| BP | GO:0030198 | extracellular matrix organization | 0.03946234 |
| BP | GO:0002673 | regulation of acute inflammatory response | 0.03946234 |
| BP | GO:0043303 | mast cell degranulation | 0.03946234 |
| BP | GO:0048146 | positive regulation of fibroblast proliferation | 0.03946234 |
| BP | GO:0043062 | extracellular structure organization | 0.04022788 |
| BP | GO:0060401 | cytosolic calcium ion transport | 0.04178876 |
| BP | GO:0001913 | T cell mediated cytotoxicity | 0.04178876 |
| BP | GO:0002279 | mast cell activation involved in immune response | 0.04178876 |
| BP | GO:0045778 | positive regulation of ossification | 0.04178876 |
| BP | GO:0046486 | glycerolipid metabolic process | 0.04178876 |
| BP | GO:0002698 | negative regulation of immune effector process | 0.04178876 |
| BP | GO:0032611 | interleukin-1 beta production | 0.04178876 |
| BP | GO:0032651 | regulation of interleukin-1 beta production | 0.04178876 |
| BP | GO:0030500 | regulation of bone mineralization | 0.04224801 |
| BP | GO:0045580 | regulation of T cell differentiation | 0.04306204 |
| BP | GO:0050730 | regulation of peptidyl-tyrosine phosphorylation | 0.04334823 |
| BP | GO:0051090 | regulation of DNA-binding transcription factor activity | 0.04344792 |
| BP | GO:0001961 | positive regulation of cytokine-mediated signaling pathway | 0.04348159 |
| BP | GO:0002448 | mast cell mediated immunity | 0.04348159 |
| BP | GO:0070231 | T cell apoptotic process | 0.04348159 |
| BP | GO:1900047 | negative regulation of hemostasis | 0.04348159 |
| BP | GO:0051279 | regulation of release of sequestered calcium ion into cytosol | 0.04348159 |
| BP | GO:0030522 | intracellular receptor signaling pathway | 0.04348159 |
| BP | GO:0045861 | negative regulation of proteolysis | 0.04348159 |
| BP | GO:1902652 | secondary alcohol metabolic process | 0.04348159 |
| BP | GO:0051896 | regulation of protein kinase B signaling | 0.04348159 |
| BP | GO:0002710 | negative regulation of T cell mediated immunity | 0.04348159 |
| BP | GO:0032682 | negative regulation of chemokine production | 0.04348159 |
| BP | GO:0042537 | benzene-containing compound metabolic process | 0.04348159 |
| BP | GO:0070977 | bone maturation | 0.04348159 |
| BP | GO:1900017 | positive regulation of cytokine production involved in inflammatory response | 0.04348159 |
| BP | GO:0002821 | positive regulation of adaptive immune response | 0.04348159 |
| BP | GO:0006887 | exocytosis | 0.04367279 |
| BP | GO:0006066 | alcohol metabolic process | 0.04441691 |
| BP | GO:0051480 | regulation of cytosolic calcium ion concentration | 0.04441691 |
| BP | GO:0034765 | regulation of ion transmembrane transport | 0.04503355 |
| BP | GO:0006636 | unsaturated fatty acid biosynthetic process | 0.04503355 |
| BP | GO:0046850 | regulation of bone remodeling | 0.04503355 |
| BP | GO:0045834 | positive regulation of lipid metabolic process | 0.04516509 |
| BP | GO:0051592 | response to calcium ion | 0.04516509 |
| BP | GO:0001894 | tissue homeostasis | 0.04518657 |
| BP | GO:0019932 | second-messenger-mediated signaling | 0.04645626 |
| BP | GO:0001959 | regulation of cytokine-mediated signaling pathway | 0.04645626 |
| BP | GO:0002227 | innate immune response in mucosa | 0.04645626 |
| BP | GO:0002825 | regulation of T-helper 1 type immune response | 0.04645626 |
| BP | GO:0034377 | plasma lipoprotein particle assembly | 0.04645626 |
| BP | GO:0046337 | phosphatidylethanolamine metabolic process | 0.04645626 |
| BP | GO:0060142 | regulation of syncytium formation by plasma membrane fusion | 0.04645626 |
| BP | GO:0071624 | positive regulation of granulocyte chemotaxis | 0.04645626 |
| BP | GO:1904062 | regulation of cation transmembrane transport | 0.04663391 |
| BP | GO:0006874 | cellular calcium ion homeostasis | 0.04663391 |
| BP | GO:0001910 | regulation of leukocyte mediated cytotoxicity | 0.04754586 |
| BP | GO:0051216 | cartilage development | 0.04823678 |
| BP | GO:0098742 | cell-cell adhesion via plasma-membrane adhesion molecules | 0.04958035 |
| BP | GO:0002707 | negative regulation of lymphocyte mediated immunity | 0.04958035 |
| CC | GO:1904724 | tertiary granule lumen | 2.14E-05 |
| CC | GO:0001533 | cornified envelope | 2.14E-05 |
| CC | GO:0009897 | external side of plasma membrane | 2.14E-05 |
| CC | GO:0034774 | secretory granule lumen | 6.58E-05 |
| CC | GO:0060205 | cytoplasmic vesicle lumen | 6.58E-05 |
| CC | GO:0031983 | vesicle lumen | 6.58E-05 |
| CC | GO:0035580 | specific granule lumen | 0.0001155 |
| CC | GO:0070820 | tertiary granule | 0.00016766 |
| CC | GO:0062023 | collagen-containing extracellular matrix | 0.00154435 |
| CC | GO:0030669 | clathrin-coated endocytic vesicle membrane | 0.00219814 |
| CC | GO:0042581 | specific granule | 0.0028527 |
| CC | GO:0045334 | clathrin-coated endocytic vesicle | 0.00797093 |
| CC | GO:0030139 | endocytic vesicle | 0.00855793 |
| CC | GO:0005775 | vacuolar lumen | 0.01933492 |
| CC | GO:0030665 | clathrin-coated vesicle membrane | 0.02706379 |
| CC | GO:0016323 | basolateral plasma membrane | 0.032968 |
| CC | GO:0035578 | azurophil granule lumen | 0.032968 |
| CC | GO:0045178 | basal part of cell | 0.03573358 |
| CC | GO:0034361 | very-low-density lipoprotein particle | 0.03841107 |
| CC | GO:0034385 | triglyceride-rich plasma lipoprotein particle | 0.03841107 |
| CC | GO:0005865 | striated muscle thin filament | 0.04837448 |
| MF | GO:0008009 | chemokine activity | 3.67E-08 |
| MF | GO:0004252 | serine-type endopeptidase activity | 3.67E-08 |
| MF | GO:0008236 | serine-type peptidase activity | 1.13E-07 |
| MF | GO:0017171 | serine hydrolase activity | 1.19E-07 |
| MF | GO:0042379 | chemokine receptor binding | 9.25E-07 |
| MF | GO:0005125 | cytokine activity | 1.52E-06 |
| MF | GO:0048018 | receptor ligand activity | 0.00010106 |
| MF | GO:0030546 | signaling receptor activator activity | 0.00011638 |
| MF | GO:0004175 | endopeptidase activity | 0.000134 |
| MF | GO:0048020 | CCR chemokine receptor binding | 0.0002937 |
| MF | GO:0048306 | calcium-dependent protein binding | 0.0002937 |
| MF | GO:0050786 | RAGE receptor binding | 0.00036169 |
| MF | GO:0004896 | cytokine receptor activity | 0.00055192 |
| MF | GO:0005126 | cytokine receptor binding | 0.00055225 |
| MF | GO:0045236 | CXCR chemokine receptor binding | 0.00383034 |
| MF | GO:0001664 | G protein-coupled receptor binding | 0.0047386 |
| MF | GO:0004222 | metalloendopeptidase activity | 0.00735285 |
| MF | GO:0140375 | immune receptor activity | 0.00853873 |
| MF | GO:0005506 | iron ion binding | 0.01144947 |
| MF | GO:0015166 | polyol transmembrane transporter activity | 0.01468793 |
| MF | GO:0061134 | peptidase regulator activity | 0.01653829 |
| MF | GO:1901618 | organic hydroxy compound transmembrane transporter activity | 0.02391296 |
| MF | GO:0004869 | cysteine-type endopeptidase inhibitor activity | 0.02934308 |
| MF | GO:0005372 | water transmembrane transporter activity | 0.02978544 |
| MF | GO:0004866 | endopeptidase inhibitor activity | 0.03010906 |
| MF | GO:0043394 | proteoglycan binding | 0.03432302 |
| MF | GO:0030414 | peptidase inhibitor activity | 0.03614671 |
| MF | GO:0008395 | steroid hydroxylase activity | 0.03900228 |
| MF | GO:0005504 | fatty acid binding | 0.04146665 |
| MF | GO:0061135 | endopeptidase regulator activity | 0.04163165 |

**Table S4.** KEGG analysis of differentially expressed genes

| ID | Description | qvalue |
| --- | --- | --- |
| hsa04061 | Viral protein interaction with cytokine and cytokine receptor | 1.07E-08 |
| hsa04060 | Cytokine-cytokine receptor interaction | 3.03E-08 |
| hsa04657 | IL-17 signaling pathway | 1.15E-06 |
| hsa05164 | Influenza A | 0.0002947 |
| hsa04062 | Chemokine signaling pathway | 0.0009557 |
| hsa04668 | TNF signaling pathway | 0.00643362 |
| hsa05160 | Hepatitis C | 0.02093557 |
| hsa04621 | NOD-like receptor signaling pathway | 0.02093557 |
| hsa05162 | Measles | 0.02405486 |
| hsa03320 | PPAR signaling pathway | 0.02774989 |
| hsa04640 | Hematopoietic cell lineage | 0.02975267 |
| hsa05171 | Coronavirus disease - COVID-19 | 0.03434538 |

**Table S5.** DO analysis of differentially expressed genes

| ID | Description | qvalue |
| --- | --- | --- |
| DOID:2348 | arteriosclerotic cardiovascular disease | 9.60E-09 |
| DOID:2349 | arteriosclerosis | 1.15E-08 |
| DOID:1936 | atherosclerosis | 1.36E-08 |
| DOID:2723 | dermatitis | 1.59E-08 |
| DOID:16 | integumentary system disease | 1.74E-08 |
| DOID:37 | skin disease | 4.66E-08 |
| DOID:3310 | atopic dermatitis | 1.83E-07 |
| DOID:0050338 | primary bacterial infectious disease | 1.87E-07 |
| DOID:403 | mouth disease | 2.09E-07 |
| DOID:104 | bacterial infectious disease | 4.89E-07 |
| DOID:3388 | periodontal disease | 9.48E-07 |
| DOID:399 | tuberculosis | 9.48E-07 |
| DOID:5295 | intestinal disease | 9.48E-07 |
| DOID:850 | lung disease | 9.48E-07 |
| DOID:75 | lymphatic system disease | 1.71E-06 |
| DOID:824 | periodontitis | 2.64E-06 |
| DOID:2237 | hepatitis | 2.64E-06 |
| DOID:2320 | obstructive lung disease | 3.72E-06 |
| DOID:3213 | demyelinating disease | 4.02E-06 |
| DOID:1091 | tooth disease | 5.77E-06 |
| DOID:2377 | multiple sclerosis | 1.04E-05 |
| DOID:552 | pneumonia | 1.83E-05 |
| DOID:9588 | encephalitis | 2.90E-05 |
| DOID:11335 | sarcoidosis | 3.63E-05 |
| DOID:1602 | lymphadenitis | 4.27E-05 |
| DOID:9942 | lymph node disease | 4.27E-05 |
| DOID:1176 | bronchial disease | 4.57E-05 |
| DOID:9008 | psoriatic arthritis | 4.63E-05 |
| DOID:2916 | hypersensitivity reaction type IV disease | 4.97E-05 |
| DOID:1883 | hepatitis C | 5.20E-05 |
| DOID:2043 | hepatitis B | 6.61E-05 |
| DOID:3908 | non-small cell lung carcinoma | 0.00013188 |
| DOID:13378 | Kawasaki disease | 0.00014496 |
| DOID:2841 | asthma | 0.00015703 |
| DOID:526 | Human immunodeficiency virus infectious disease | 0.00017333 |
| DOID:0060005 | autoimmune disease of endocrine system | 0.00021739 |
| DOID:0050589 | inflammatory bowel disease | 0.00029303 |
| DOID:120 | female reproductive organ cancer | 0.00030293 |
| DOID:1398 | parasitic infectious disease | 0.00048219 |
| DOID:11729 | Lyme disease | 0.00048219 |
| DOID:3082 | interstitial lung disease | 0.00048219 |
| DOID:12361 | Graves' disease | 0.00050022 |
| DOID:28 | endocrine system disease | 0.0005612 |
| DOID:4989 | pancreatitis | 0.00062076 |
| DOID:0060180 | colitis | 0.00078439 |
| DOID:8577 | ulcerative colitis | 0.00078439 |
| DOID:3770 | pulmonary fibrosis | 0.00094813 |
| DOID:3393 | coronary artery disease | 0.00127315 |
| DOID:13406 | pulmonary sarcoidosis | 0.00131045 |
| DOID:0050700 | cardiomyopathy | 0.00144145 |
| DOID:2797 | idiopathic interstitial pneumonia | 0.00148528 |
| DOID:26 | pancreas disease | 0.0015307 |
| DOID:9352 | type 2 diabetes mellitus | 0.00159276 |
| DOID:8469 | influenza | 0.00164329 |
| DOID:7998 | hyperthyroidism | 0.00180723 |
| DOID:3083 | chronic obstructive pulmonary disease | 0.00192771 |
| DOID:5082 | liver cirrhosis | 0.00192771 |
| DOID:345 | uterine disease | 0.00193686 |
| DOID:7693 | abdominal aortic aneurysm | 0.00213499 |
| DOID:4450 | renal cell carcinoma | 0.00213499 |
| DOID:854 | collagen disease | 0.00213499 |
| DOID:50 | thyroid gland disease | 0.00221676 |
| DOID:1107 | esophageal carcinoma | 0.00296034 |
| DOID:2893 | cervix carcinoma | 0.00298754 |
| DOID:5844 | myocardial infarction | 0.00302786 |
| DOID:4362 | cervical cancer | 0.00302786 |
| DOID:5041 | esophageal cancer | 0.00302786 |
| DOID:1575 | rheumatic disease | 0.00302786 |
| DOID:418 | systemic scleroderma | 0.00302786 |
| DOID:419 | scleroderma | 0.00302786 |
| DOID:3459 | breast carcinoma | 0.00330842 |
| DOID:1564 | fungal infectious disease | 0.00545793 |
| DOID:1273 | respiratory syncytial virus infectious disease | 0.00545793 |
| DOID:3007 | breast ductal carcinoma | 0.00546794 |
| DOID:0060036 | intrinsic cardiomyopathy | 0.00546794 |
| DOID:3627 | aortic aneurysm | 0.00592061 |
| DOID:520 | aortic disease | 0.00645488 |
| DOID:4451 | renal carcinoma | 0.00660202 |
| DOID:4074 | pancreas adenocarcinoma | 0.00660202 |
| DOID:10124 | corneal disease | 0.00660202 |
| DOID:3070 | malignant glioma | 0.00660202 |
| DOID:4606 | bile duct cancer | 0.00660202 |
| DOID:4897 | bile duct carcinoma | 0.00660202 |
| DOID:9744 | type 1 diabetes mellitus | 0.00660202 |
| DOID:3480 | uveal disease | 0.00675738 |
| DOID:3748 | esophagus squamous cell carcinoma | 0.00700806 |
| DOID:2986 | IgA glomerulonephritis | 0.00715324 |
| DOID:1168 | familial hyperlipidemia | 0.00742034 |
| DOID:2394 | ovarian cancer | 0.00814773 |
| DOID:11934 | head and neck cancer | 0.00869739 |
| DOID:229 | female reproductive system disease | 0.00929143 |
| DOID:8778 | Crohn's disease | 0.00969786 |
| DOID:2789 | parasitic protozoa infectious disease | 0.00969878 |
| DOID:1793 | pancreatic cancer | 0.00986502 |
| DOID:4896 | bile duct adenocarcinoma | 0.00996043 |
| DOID:4947 | cholangiocarcinoma | 0.00996043 |
| DOID:654 | overnutrition | 0.01059261 |
| DOID:9455 | lipid storage disease | 0.01094401 |
| DOID:9415 | allergic asthma | 0.01104886 |
| DOID:4766 | embryoma | 0.01146538 |
| DOID:2089 | constipation | 0.01146538 |
| DOID:4988 | alcoholic pancreatitis | 0.01146538 |
| DOID:9779 | bowel dysfunction | 0.01146538 |
| DOID:7166 | thyroiditis | 0.0115365 |
| DOID:635 | acquired immunodeficiency syndrome | 0.0115365 |
| DOID:4607 | biliary tract cancer | 0.01168445 |
| DOID:3405 | histiocytosis | 0.01168445 |
| DOID:3146 | lipid metabolism disorder | 0.01259393 |
| DOID:9261 | nasopharynx carcinoma | 0.01267221 |
| DOID:0060049 | autoimmune disease of urogenital tract | 0.01267221 |
| DOID:12236 | primary biliary cirrhosis | 0.01267221 |
| DOID:3744 | cervical squamous cell carcinoma | 0.01267221 |
| DOID:12930 | dilated cardiomyopathy | 0.0127911 |
| DOID:6050 | esophageal disease | 0.0127911 |
| DOID:6543 | acne | 0.0132541 |
| DOID:9098 | sebaceous gland disease | 0.0132541 |
| DOID:3996 | urinary system cancer | 0.01330108 |
| DOID:74 | hematopoietic system disease | 0.01394847 |
| DOID:9452 | fatty liver disease | 0.01394847 |
| DOID:374 | nutrition disease | 0.01440355 |
| DOID:0060119 | pharynx cancer | 0.0145756 |
| DOID:1542 | head and neck carcinoma | 0.0145756 |
| DOID:9970 | obesity | 0.01477996 |
| DOID:8398 | osteoarthritis | 0.01502082 |
| DOID:2462 | retinal vascular disease | 0.01512849 |
| DOID:8947 | diabetic retinopathy | 0.01512849 |
| DOID:12571 | phacogenic glaucoma | 0.01513873 |
| DOID:12895 | keratoconjunctivitis sicca | 0.01513873 |
| DOID:13641 | exfoliation syndrome | 0.01513873 |
| DOID:9368 | keratoconjunctivitis | 0.01513873 |
| DOID:2600 | laryngeal carcinoma | 0.01560287 |
| DOID:3978 | extrinsic cardiomyopathy | 0.01560287 |
| DOID:820 | myocarditis | 0.01560287 |
| DOID:688 | embryonal cancer | 0.0163416 |
| DOID:2921 | glomerulonephritis | 0.01636263 |
| DOID:865 | vasculitis | 0.016985 |
| DOID:10140 | dry eye syndrome | 0.01779789 |
| DOID:1400 | lacrimal apparatus disease | 0.01779789 |
| DOID:633 | myositis | 0.01810298 |
| DOID:9074 | systemic lupus erythematosus | 0.01810298 |
| DOID:10952 | nephritis | 0.01836118 |
| DOID:1555 | urticaria | 0.01887763 |
| DOID:12365 | malaria | 0.01895992 |
| DOID:263 | kidney cancer | 0.01901204 |
| DOID:2994 | germ cell cancer | 0.01947981 |
| DOID:3211 | lysosomal storage disease | 0.02035168 |
| DOID:5520 | head and neck squamous cell carcinoma | 0.02035168 |
| DOID:3910 | lung adenocarcinoma | 0.02385111 |
| DOID:8857 | lupus erythematosus | 0.02501673 |
| DOID:289 | endometriosis | 0.02640516 |
| DOID:10283 | prostate cancer | 0.02742325 |
| DOID:299 | adenocarcinoma | 0.02787168 |
| DOID:1206 | Rett syndrome | 0.02791452 |
| DOID:240 | iris disease | 0.02791452 |
| DOID:9675 | pulmonary emphysema | 0.02791452 |
| DOID:3069 | astrocytoma | 0.02839406 |
| DOID:3355 | fibrosarcoma | 0.03130605 |
| DOID:9408 | acute myocardial infarction | 0.03145998 |
| DOID:9500 | leukocyte disease | 0.03145998 |
| DOID:4248 | coronary stenosis | 0.03182353 |
| DOID:3856 | male reproductive organ cancer | 0.03271431 |
| DOID:10223 | dermatomyositis | 0.03327367 |
| DOID:13250 | diarrhea | 0.03327367 |
| DOID:4251 | conjunctival disease | 0.03327367 |
| DOID:0060100 | musculoskeletal system cancer | 0.03503669 |
| DOID:12716 | newborn respiratory distress syndrome | 0.03592602 |
| DOID:2596 | larynx cancer | 0.03592602 |
| DOID:3683 | lung benign neoplasm | 0.03592602 |
| DOID:1115 | sarcoma | 0.03696191 |
| DOID:381 | arthropathy | 0.0386301 |
| DOID:0050621 | respiratory system benign neoplasm | 0.03890011 |
| DOID:11162 | respiratory failure | 0.03907676 |
| DOID:438 | autoimmune disease of the nervous system | 0.03907676 |
| DOID:655 | inherited metabolic disorder | 0.03907676 |
| DOID:3498 | pancreatic ductal adenocarcinoma | 0.03953022 |
| DOID:18 | urinary system disease | 0.04064153 |
| DOID:10113 | trypanosomiasis | 0.04377175 |
| DOID:8283 | peritonitis | 0.04377175 |
| DOID:0050904 | salivary gland carcinoma | 0.04377175 |
| DOID:8850 | salivary gland cancer | 0.04663599 |
| DOID:423 | myopathy | 0.04723885 |
| DOID:66 | muscle tissue disease | 0.04723885 |
| DOID:2473 | opportunistic mycosis | 0.04772708 |
| DOID:0060071 | pre-malignant neoplasm | 0.04804627 |
| DOID:3369 | peripheral primitive neuroectodermal tumor | 0.04804627 |
| DOID:1428 | endocrine pancreas disease | 0.04804627 |
| DOID:10652 | Alzheimer's disease | 0.04804627 |
| DOID:2151 | malignant ovarian surface epithelial-stromal neoplasm | 0.04804627 |
| DOID:2152 | ovary epithelial cancer | 0.04804627 |
| DOID:4001 | ovarian carcinoma | 0.04804627 |
| DOID:557 | kidney disease | 0.049029 |
| DOID:4866 | salivary gland adenoid cystic carcinoma | 0.04952028 |

**Table S6.** The weighted values in the trained neural network

| Neural Link | Weighted Value |
| --- | --- |
| Intercept.to.1layhid1 | -2.2762975 |
| BTC.to.1layhid1 | -21.371404 |
| PPP4R1.to.1layhid1 | 3.41376173 |
| SERPINB4.to.1layhid1 | -17.586226 |
| S100A7.to.1layhid1 | -19.196819 |
| S100A9.to.1layhid1 | -17.898785 |
| GALNT6.to.1layhid1 | 0.63382358 |
| Intercept.to.1layhid2 | 2.12377447 |
| BTC.to.1layhid2 | 9.97362158 |
| PPP4R1.to.1layhid2 | 1.45802283 |
| SERPINB4.to.1layhid2 | -0.9386225 |
| S100A7.to.1layhid2 | 0.95343847 |
| S100A9.to.1layhid2 | -0.9047703 |
| GALNT6.to.1layhid2 | -0.9045004 |
| Intercept.to.1layhid3 | 0.13193526 |
| BTC.to.1layhid3 | 0.96509672 |
| PPP4R1.to.1layhid3 | -17.502003 |
| SERPINB4.to.1layhid3 | -17.699035 |
| S100A7.to.1layhid3 | -18.533714 |
| S100A9.to.1layhid3 | -15.619123 |
| GALNT6.to.1layhid3 | -0.8706702 |
| Intercept.to.1layhid4 | -1.5840081 |
| BTC.to.1layhid4 | -20.886021 |
| PPP4R1.to.1layhid4 | -2.0875638 |
| SERPINB4.to.1layhid4 | 1.21284453 |
| S100A7.to.1layhid4 | -1.7766953 |
| S100A9.to.1layhid4 | 2.03325539 |
| GALNT6.to.1layhid4 | 1.19603959 |
| Intercept.to.Normal | 0.24206428 |
| 1layhid1.to.Normal | -1.2584605 |
| 1layhid2.to.Normal | 0.74432078 |
| 1layhid3.to.Normal | -1.3054917 |
| 1layhid4.to.Normal | -0.4615753 |
| Intercept.to.AD | -0.4866204 |
| 1layhid1.to.AD | 1.28502307 |
| 1layhid2.to.AD | 0.50051816 |
| 1layhid3.to.AD | 1.3258898 |
| 1layhid4.to.AD | 1.1552478 |
